# A transient decrease in night-to-night variability of REM sleep before myocardial infarction

**DOI:** 10.64898/2026.10.26.26366566

**Authors:** Edo Ikurumi, Junichi Sugita, Kensuke Ueno, Naoto Setoguchi, Hiroshi Seno, Ryoko Uchida, Yiyi Yang, Zheng Liu, Sumie Miyamoto, Kantaro Fujiwara, Kazuyuki Aihara, Katsuhito Fujiu

## Abstract

**Background:** Myocardial infarction (MI) outcomes depend on time to treatment, and fatigue and sleep disturbance are often reported in the weeks before the event. Whether wearable-measured sleep changes within individuals before MI, and whether such change is specific to MI, is unknown.

**Methods:** In the All of Us Research Program, adults with a first MI code and at least 180 valid Fitbit sleep nights in the preceding 360 days were matched to up to 10 controls on sex, comorbidities, sleep apnea, calendar time and a propensity score. The full matched cohort of all cases was the primary cohort, and cases with diabetes and/or hypertension formed a subgroup. The main endpoints were the within-person change in night-to-night variability (SD, RMSSD, CV) of the REM-sleep fraction at days 30–16 relative to each person’s baseline (days 360–121) and from days 30–16 to days 15–1, tested against 10,000 randomly placed index dates with allowance for the choice of window (max-T). The same pipeline was applied to cases without recorded symptoms or treatment, to an index date at the first angina code, to a cohort including heart failure and to ischaemic stroke and admissions for heart failure, pneumonia, fracture or appendicitis and any cause, and the analysis was adjusted for nightly heart rate, wear and device.

**Results:** The full matched cohort comprised 45 cases and 450 controls. REM-fraction variability changed transiently before MI, decreasing at days 30–16 and returning towards baseline over days 15–1; allowing for the choice of window, this decrease-then-return pattern was rarely matched at 10,000 randomly placed index dates (max-T p<0.001). Its components were the decrease in SD at days 30–16 (Cliff’s δ −0.30, p=0.004; OR per SD 0.53, 95% CI 0.35–0.80; max-T p=0.0034) and the return in RMSSD over days 15–1 (δ +0.36, p<0.001; max-T p=0.0035); day by day, the decrease was not detected as a continuous change (cluster permutation test p=0.58). The decrease in SD was not seen at the same time before admissions for pneumonia (69 cases), fracture or appendicitis (130) or any cause (859) and was larger before MI than before each of them (difference in δ −0.25 to −0.34), whereas the rise over days 15–1 was shared with pneumonia; it was of similar size in the subgroup with diabetes and/or hypertension, in cases without recorded symptoms or treatment, with the index date at the first angina code, when people with heart failure were included and after adjustment for nightly heart rate, wear and device. Scoring each person against their own baseline discriminated cases from controls with AUCs of up to 0.66, probably an optimistic estimate because the decrease in RMSSD was attenuated in held-out halves of the matched sets.

**Conclusions:** In adults who use consumer wearables, a first MI was preceded by a transient change in night-to-night REM-sleep variability, a decrease at days 30–16 and a return over the following two weeks. The decrease was not seen at the same time before other acute admissions, did not depend on recorded symptoms or treatment and was detectable against each person’s own baseline with simple measures. The timing is an exploratory finding, to be confirmed in new cases.

## Introduction

In the United States, a myocardial infarction (MI) occurs about every 40 seconds, and ischaemic heart disease caused about 9 million deaths worldwide in 2021 ^1^. Outcomes depend on time to treatment: each 30 minutes of delay to primary angioplasty was associated with a 7.5% relative increase in 1-year mortality ^2^. In a Swedish registry of 89,155 patients with ST-elevation MI followed over 20 years, the median pre-hospital delay was 150 minutes; each hour of delay independently increased 14-day and 1-year mortality, and delay did not decline over the two decades ^3^. In a Japanese registry of 4,625 patients, 63.3% arrived 2 hours or more after symptom onset, and older age, diabetes and night-time onset were associated with longer delay ^4^.

Many patients report symptoms before their MI. In a study of 515 women, 95% reported new or different symptoms more than a month before the event, most often unusual fatigue (70%) and sleep disturbance (48%) ^5^. In a cross-sectional study of 242 patients, 41.3% reported warning symptoms, including unusual fatigue (23%) and sleep disturbance (22%) ^6^.

Sleep is closely linked to cardiovascular health. In a dose-response meta-analysis of 71 cohorts comprising 3.8 million participants, sleep duration showed a U-shaped association with cardio-cerebrovascular disease, with the lowest risk at about 7.5 hours per night ^7^. In the All of Us Research Program, sleep measured with consumer wearables over a median of 4.5 years in 6,785 participants was associated with a range of incident chronic diseases: irregular sleep with obesity, hyperlipidaemia, hypertension, major depressive disorder and generalised anxiety disorder, and REM and deep sleep inversely with atrial fibrillation ^8^.

Among US mobile phone users, 28% reported using an app to track their sleep ^9^. Against polysomnography, a current Fitbit device showed no significant bias in total sleep time, while agreement for sleep stages was moderate (four-stage agreement 70.9%, κ 0.55) ^10^. Continuous wearable monitoring relative to a person’s own baseline has anticipated acute events in other conditions: in patients with heart failure, a multisensor patch predicted rehospitalisation with 76–88% sensitivity and 85% specificity at a median of 6.5 days before admission ^11^, and a wrist-worn device detected SARS-CoV-2 infection earlier than symptom reporting, although with low specificity ^12^. To our knowledge, within-person changes in wearable-measured sleep in the weeks before MI have not been examined; existing wearable studies have related habitual sleep to long-term disease risk ^8^ or detected short-term change before other acute events ^11,12^.

In the All of Us Research Program, heart-rate and step-count features from Fitbit devices predicted incident cardiovascular disease (area under the curve 0.80) and all-cause hospitalisation in 14,157 participants ^13^, and in the period from 90 days before to 30 days after a diagnosis of incident heart failure, the percentage of REM sleep decreased and that of light sleep increased, whereas matched controls did not change^14^.

Using long-term Fitbit sleep data linked to electronic health records in the All of Us Research Program, we examined within-person changes in night-to-night variability of REM sleep in the weeks before incident MI, compared with matched controls, and tested whether the changes were specific to MI by applying the same analysis to other acute events.

## Methods

### Data source

We used the All of Us Research Program Registered Tier Dataset v9 (curated data repository R2025Q4R6), which links Fitbit data to electronic health records (EHR). See Declarations for ethics and data availability.

### Cases

The index date was the first EHR code for acute or unspecified MI (ICD-10-CM I21.0– I21.4, I21.9, I21.A9; ICD-9-CM 410.x0/410.x1). Participants were excluded if a code for prior MI (I25.2, 412), post-MI or subsequent MI (I22–I23, 410.x2, 411.0, 429.7x) or type 2 MI (I21.A1) was recorded before the index date. Cases were required to have age and sex recorded, at least 180 valid sleep nights in the 360 days before the index date, a body mass index (BMI; latest value within 3 years) and baseline sleep available, and no atrial fibrillation or flutter (I48; 427.31/427.32) or heart-failure (I50.x; 428.x) code before the index date. Atrial fibrillation and heart failure were excluded because they disturb sleep architecture and heart-rate-based sleep staging; a sensitivity analysis included heart failure (Secondary analyses).

### Sleep data

A valid night was a main-sleep record with sleep stages and at least 180 minutes asleep; nights with zero REM sleep (under 1% of nights) were retained with a REM fraction of 0. The REM fraction was REM minutes divided by minutes asleep. Deep and light fractions, total sleep time, the wake fraction (minutes awake divided by time in bed) and sleep efficiency were derived for exploratory analyses. The baseline period was days 360–121 before the index date; at least 30 valid baseline nights were required.

### Controls and matching

Candidate controls had no MI-class code and a 360-day window of Fitbit data ending within ±365 days of the case’s index date. For each control, one candidate reference date was set in every calendar week, on a pseudo-random day of that week, irrespective of whether sleep was recorded that night, so that the window end did not depend on device wear; for each case, the candidate nearest to the case’s index date was used. Controls were matched exactly on sex, diabetes (E08–E13; 250), hypertension (I10–I16; 401–405) and sleep apnea (G47.3; 327.2, 780.51/53/57) status, and by a propensity score for age, BMI and baseline sleep duration within a caliper of 0.5 SD of the logit ^15^, taking 5–10 controls per case, nearest first, without replacement. The same exclusions as for cases applied. The full matched cohort was the primary cohort; matched sets whose case had diabetes and/or hypertension (subgroup) and sets without either condition (sensitivity) were also analysed. The number of cases in the subgroup is not reported because, together with the full matched cohort, it would reveal a count of 20 or fewer (the sets without diabetes or hypertension).

### Endpoints

For each person, night-to-night variability of the REM fraction was summarised in 15-night windows by the SD, the root mean square of successive differences between consecutive calendar nights that were both valid (RMSSD) and the coefficient of variation (CV). Endpoint 1 was the change at days 30–16 relative to the person’s baseline: the median, over all same-length baseline windows, of the difference between the target window and each baseline window. Endpoint 2 was the change from days 30– 16 to days 15–1, and endpoint 3 the change at days 15–1 relative to baseline. Windows required at least 60% valid nights and were compared with equal numbers of valid nights. SD and RMSSD changes were divided by the person’s nightly scale (1.4826 × median absolute deviation of the baseline); CV changes were used as plain differences. SD and RMSSD are related but capture different timescales: SD reflects overall spread within the window, including slow drifts, whereas RMSSD reflects only differences between consecutive nights (RMSSD² ≈ 2·SD²·(1 − lag-1 autocorrelation)).

Endpoint selection. The direction and approximate timing of the change were first seen in a previous All of Us data release (CDR v8), which overlaps with the present cohort and so does not provide independent validation. In an analysis reported on 7 August 2026, the 10–30 days before MI (near period) were compared within each case with the 20 days with the most data earlier than 50 days before MI (far period), each period with at least 5 days of data; among activity, heart-rate and sleep features, the SD of daily REM minutes (minute_rem_std) separated the two periods most strongly and was lower in the near period in every case (Wilcoxon p<0.01; the number of cases is not shown, 20 or fewer). Variability of the nightly REM fraction was carried forward from analyses of that release: across them, measures of night-to-night variability (SD, RMSSD, CV) consistently separated MI cases from controls, whereas the mean REM fraction did not. The within-person method (each window scored against the person’s own same-length baseline windows, with nights equalised) and the 15-night window length were also fixed on v8. In the present data, the decrease seen in v8 was refined to the windows of days 30–16 and 15–1 after an exploratory analysis. We therefore refer to them as main rather than primary endpoints, treat them as exploratory and tested them against randomly placed index dates (Robustness checks).

### Secondary analyses

#### Cases without recorded symptoms or treatment

A case was considered symptomatic if, in days 90–1 before the index date, a beta-blocker, nitrate, antiplatelet or calcium-channel blocker was newly started (no record of the same class in the preceding 365 days) or an angina (I20.x) or chest-pain (R07.x) code was recorded, and asymptomatic otherwise. Matched sets with an asymptomatic case were analysed with the statistics of the main endpoints.

#### Other acute events

To test whether the changes were specific to MI, the pipeline (data requirements, matching and endpoints) was applied unchanged to ischaemic stroke and to admissions for heart failure, pneumonia, fracture or appendicitis (an event without a physiological prodrome) and any cause, each with its own matched controls. For stroke, the index date was the first acute ischaemic stroke code (I63; 433.x1, 434.x1) within an acute visit (inpatient, emergency or intensive care) containing that day or starting up to 3 days before it; earlier stroke or stroke-history codes, lacunar or cerebral venous codes in the index episode, MI before the index date and atrial fibrillation or flutter before or within 365 days after it excluded a person. For heart failure (I50; 428), pneumonia (J12–J18; 480–486) and fracture or appendicitis (S02, S12, S22, S32, S42, S52, S62, S72, S82, S92, K35; 800–829, 540), the person’s first code of the group had to lie in such an acute visit, and the index date was the start of that visit; for any admission, it was the first inpatient admission. For pneumonia and any admission, a person was excluded if an acute MI code (I21–I22; 410) was recorded during the index admission, from its start to its discharge (or to the first pneumonia code, if later). In the admission groups, an MI or stroke code before the index date and, except for heart failure, atrial fibrillation or flutter excluded a person. Controls had no code of the group before the reference date or within 365 days after it. A heart-failure code before the index or reference date excluded cases and controls in every group, as for MI. Power to detect the effect seen in MI (δ −0.30) was estimated with the Noether approximation ^16^. MI was compared with each admission group directly by the difference in Cliff’s δ of endpoint 1 (MI minus group), with a 95% CI from 2,000 bootstrap resamples in which the matched sets of MI and of the group were resampled independently. Persons included in both the MI analysis and a comparison analysis, as a case or a control, were kept in both (43 for pneumonia, 53 for fracture or appendicitis and 213 for any admission).

#### Heart failure

The whole analysis, including extraction and matching, was repeated in a cohort in which heart failure neither excluded participants nor was used for matching; the size of this cohort is not reported because, together with the main cohort, it would reveal the number of people with heart failure. In that cohort, controls with and without a heart-failure code were compared, the per-person scores of MI cases were compared against each and the conditional logistic regression was repeated with heart-failure status as a covariate.

Nightly heart rate, wear and device. Minute-level heart rate (25–220 beats per minute) within the main-sleep segments gave, for each valid night, the mean heart rate and the share of main-sleep minutes with a heart-rate value (wear fraction). The changes at days 30–16 of the nightly mean heart rate and of its 15-night SD, computed with the same within-person method as the REM endpoint, were always included as covariates. The valid-night fraction and the wear fraction at days 30– 16 and the Fitbit device (the wrist device recorded on the valid nights at days 30– 16 or the following day, scales and phone tracking excluded, grouped as Charge, Inspire, Versa, Sense, other or no record; per person, the family with the most such nights, ties going to the most recent) were compared between cases and controls by the standardised mean difference with controls weighted by 1/(controls in the set), and those of 0.1 or more in absolute value were added, the largest first, up to four variables in all; for the device the largest absolute SMD over its categories was used, and it was entered as indicator variables against the most common family. A missing wear fraction was set to the mean. The conditional logistic regression was fitted with and without these covariates in the people with both heart-rate covariates.

#### Index date at the first angina code

For cases whose first-ever angina code (I20.x, including unstable angina) lay in days 90–1 before the MI index date, the whole matched set was re-indexed: the case’s index date was set to that code and the controls’ reference dates were moved back by the same number of days, and all windows were recomputed. The first angina or chest-pain code (I20.x or R07.x) was used as a secondary definition, and the analysis was repeated with these sets removed.

#### Composite indices

From the per-person z scores, we formed the dip (−z of days 30– 16), the rise over days 15–1 (z of days 15–1 minus z of days 30–16, or the z score of the change from days 30–16 to days 15–1 against the person’s changes between adjacent baseline windows), their conjunction (the minimum of dip and rise) and a flag (dip z ≤ −1 and rise ≥ +1), and computed the AUC for MI cases against their own controls and against the cases of each other event group. Secondary analyses were not adjusted for multiplicity.

## Statistical analysis

Changes were compared between cases and controls with Cliff’s δ and the Mann– Whitney test ^17^, conditional logistic regression within matched sets (odds ratio per SD) ^18^ and a within-set permutation test (1,000 permutations). Each group’s change from zero was summarised by the matched-pairs rank-biserial correlation with the Wilcoxon signed-rank test ^19^. To allow for residual imbalance after matching, the conditional logistic regression was repeated with age, BMI and baseline sleep duration (standardised) as covariates.

### Per-person detection

Each person’s change was expressed as a z score against their own distribution of baseline windows (median and 1.4826 × MAD). We report the AUC, the odds ratio for being flagged (|z| ≥ 1.645) and, for continuous monitoring, the number of false alarms per control-year over days 120–1.

### Exploratory segment analysis

For every sleep measure, nine window features (mean, median, SD, CV, median absolute deviation, minimum, maximum, RMSSD and lag-1 autocorrelation) were computed in disjoint 15-, 30-and 60-night segments over days 120–1, each compared with same-length baseline windows and with the preceding segment, using the same statistics as the main endpoints.

### Exploratory analyses

Bubble entropy ^20^, distribution entropy ^21^ (pairs at lags ≤ 10 nights), local variation (Lv) ^22^ over 14 and 30 nights, Bayesian online changepoint detection ^23^ and a Kalman-filtered time-varying vector autoregression ^24^ were computed for each sleep measure and compared as above. Person-level logistic regression models combining endpoint features were evaluated by leave-one-matched-set-out cross-validation, and day-level classifiers (logistic regression and random forest) by 5-fold cross-validation grouped by matched set. Exploratory p values are not adjusted for multiplicity.

### Robustness checks

To test whether a comparable decrease and increase could arise at any time, each matched set (the case and its controls together) was assigned a random earlier index date, drawn from the offsets of 60–354 days (7-day steps) at which its case met the same data requirement as at the real index date and had an endpoint-1 value; a control was included at that date only if the preceding 360 days held at least 180 valid nights, and sets whose case had no such offset were not used (the observed values were recomputed in the same sets). Endpoint 1, endpoint 2 and a combined decrease-then-increase statistic, min(−δ1, δ2), were recomputed with the same pipeline in 10,000 draws, a number chosen for precision. Single-window p values were two-sided (twice the smaller tail of the draws). To allow for the window having been chosen after the fact, the same statistics were computed at each of the eight 15-night windows in days 120–1 in every draw; each window’s statistic was standardised by the mean and SD of its own random-date distribution, and the observed standardised value at the paper’s window was compared with the largest across the windows in each draw; two-sided, the largest absolute value was used for endpoints 1 and 2 and, for the combined statistic (seven window positions), the larger of it and its mirror, min(δ1, −δ2) (max-T). In a cluster permutation test, in the same matched sets, the 14-night rolling SD of Figure 1b was compared between cases and controls on each of days 120–1 on which both groups had more than 20 people with a value (Mann–Whitney); runs of consecutive days with p<0.05 and the same sign formed clusters, and the mass of the largest decrease cluster overlapping days 30–16 (the sum of the daily differences) was compared with the largest absolute cluster mass over days 120–1 in each of the same 10,000 draws. As secondary analyses, the cluster test was repeated one-sided (decrease clusters against the largest decrease-cluster mass) and with a forming threshold of p<0.01, and max-T was computed one-sided and over the three windows of days 45–31, 30–16 and 15–1; the one-sided versions and the three-window range rest on the decrease seen in the previous data release (Endpoint selection). In a split-half check, the matched sets were divided at random into two halves 1,000 times; in one half the window with the largest |δ| among the eight was chosen, and δ was measured at that window in the other half, with its sign reversed when the chosen change was an increase, so that a negative value means the chosen change repeated. The check was repeated with a shorter baseline (days 240–121, random dates 60–116 days earlier), applied to real and random dates alike. Endpoints 1–3 were re-estimated in persons with all three available. For the index date, inpatient and emergency visits within 30 days of the first MI code were identified. Where the visit began before the code date and the diagnoses recorded before the code indicated an MI presentation (chest pain, angina, tachycardia or palpitations, dyspnoea, pain, hypertensive urgency), the index date was moved to the visit start; cases with a separate acute illness recorded first (stroke, thyrotoxicosis) were kept at the code date. In addition, all windows were moved 3 and 5 days earlier, leaving out the final nights before the index date. Missing nights were classified as no main-sleep record, under 3 hours asleep, no sleep stages or valid with zero REM sleep (retained as a REM fraction of 0).

**Figure 1.**
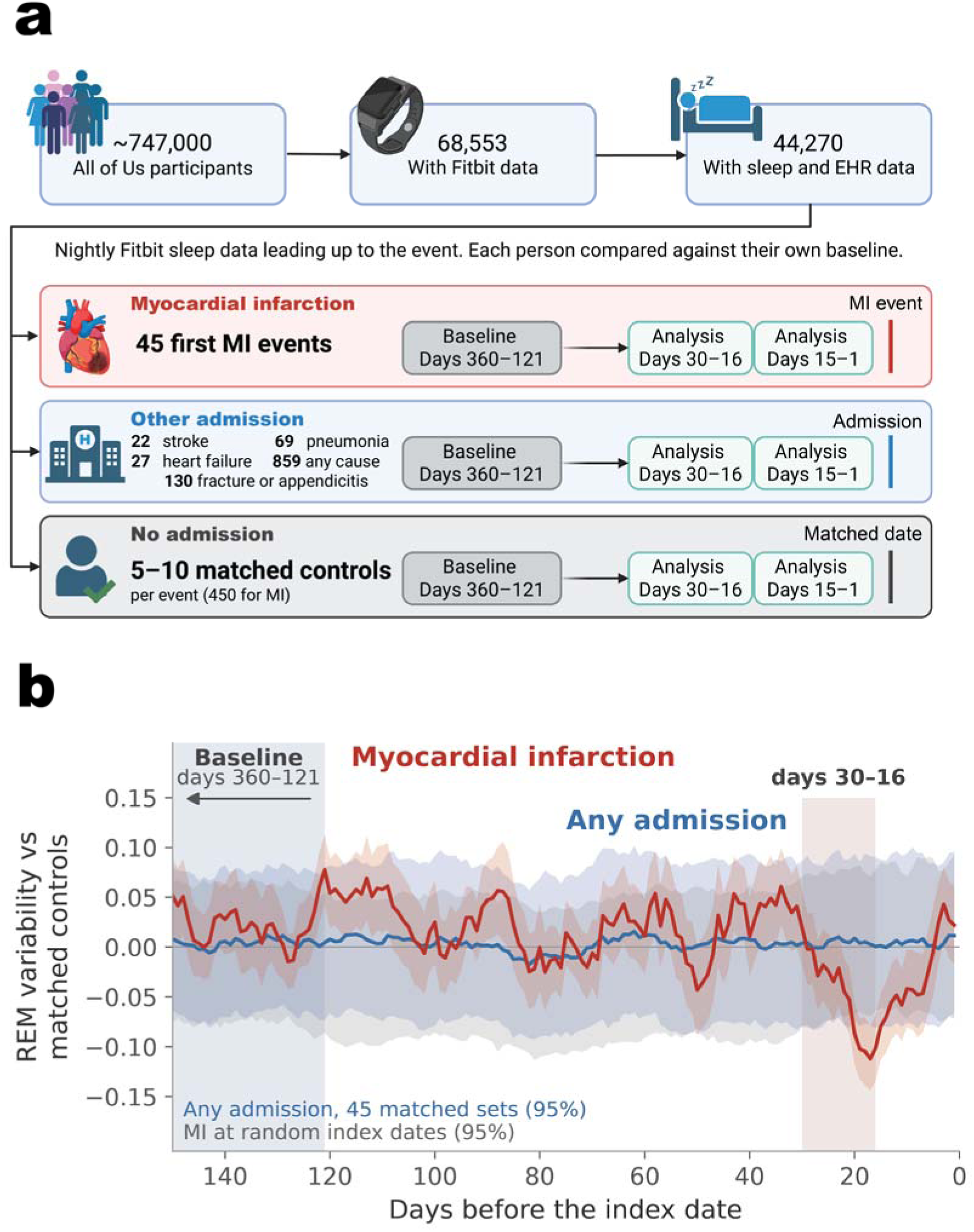
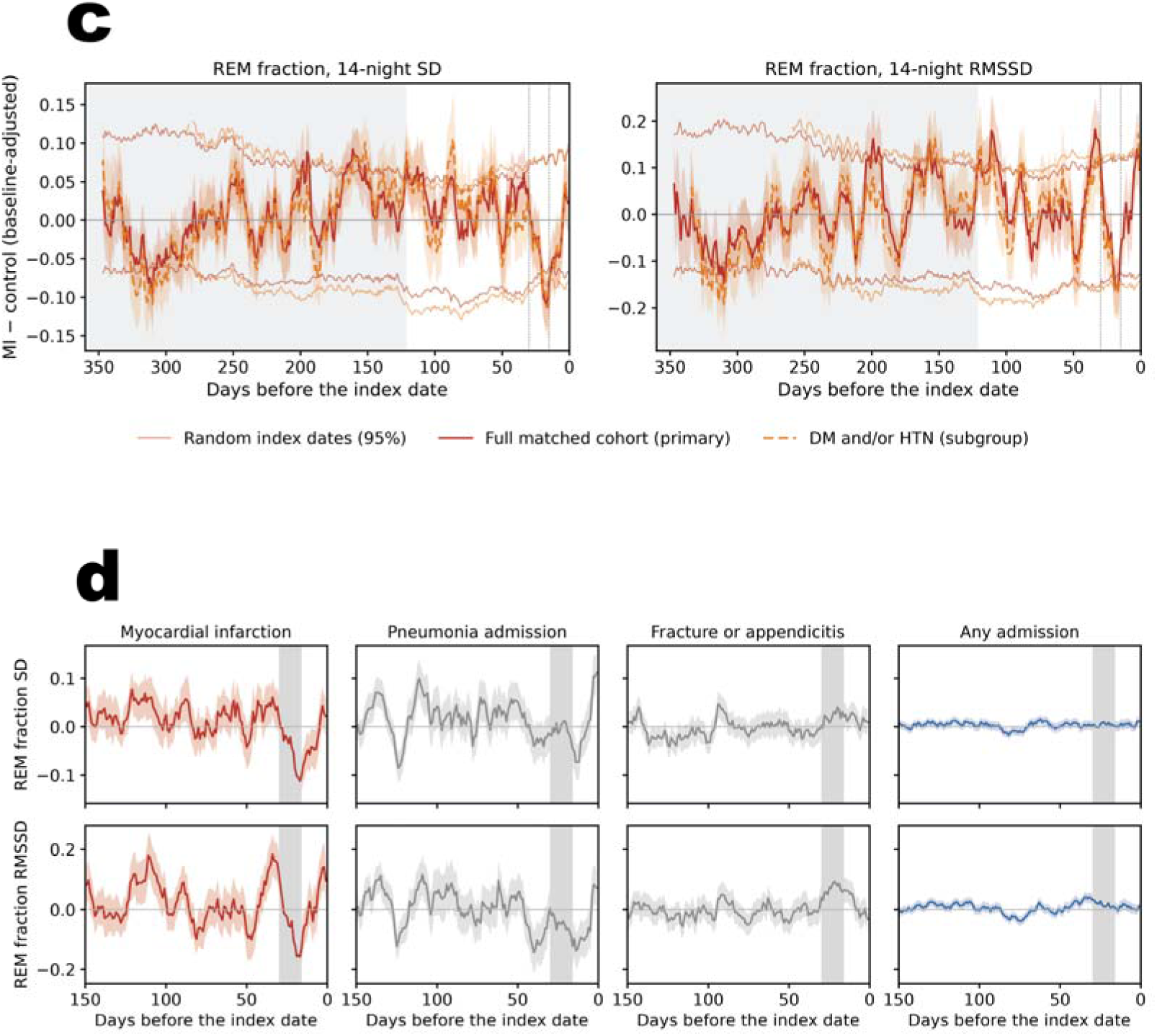
Study design and REM-fraction variability before MI and before other acute events. (a) Participants in All of Us Registered Tier Dataset v9 with Fitbit sleep data and linked EHR; first MI and admissions for ischaemic stroke, heart failure, pneumonia, fracture or appendicitis and any cause, each with 5–10 matched controls per case. Nightly sleep at days 30–16 and 15–1 before the index date was compared with each person’s own baseline (days 360–121). Created in https://BioRender.com. (b) Difference between cases and matched controls (MI ±SE) in 14-night rolling SD of the REM fraction, adjusted to each person’s baseline, before MI and before admission for any cause (full matched cohort), over days 150–1. Grey: baseline period; shaded: days 30–16; grey band: central 95% of the MI curve over 10,000 draws in which the index date of each matched set was moved to a random earlier date 60–354 days before (7-day steps) at which its case met the same data requirement, drawn only on days on which at least 1,000 draws had a value; blue band: central 95% of the curve for any admission over 2,000 draws of 45 of its matched sets, drawn at random without replacement. (c) Difference between cases and matched controls (±SE) in 14-night rolling SD (left) and RMSSD (right) of the REM fraction, adjusted to each person’s baseline, in the full matched cohort (primary) and the subgroup with diabetes and/or hypertension. Grey: baseline period; dotted lines: days 30 and 15; thin lines: central 95% of the same curve over the 10,000 draws with randomly placed index dates described in (b). (d) The same difference before MI and before admissions for pneumonia, fracture or appendicitis and any cause (full matched cohort; pneumonia and any admission exclude admissions with an acute MI code), for SD (top) and RMSSD (bottom), over days 150–1. Shaded: days 30–16. Ischaemic stroke and heart-failure admission are not shown because most days had 20 or fewer people with data. Days on which either group had 20 or fewer people with data are omitted. Rolling 14-night windows are shown for display; the endpoints use 15-night windows (Methods).

### Disclosure control

In accordance with the All of Us Data and Statistics Dissemination Policy, no count of 1–20, or value from which one can be derived, is reported; such cells are shown as masked. Analyses used Python 3.11.8 with NumPy 1.26.4, pandas 2.1.4, SciPy 1.11.4, scikit-learn 1.3.2, Matplotlib 3.8.4 and pandas-gbq 0.35.2.

## Results

### Study population

The study design is shown in Figure 1a and the participant flow in Figure S1. Of 790 participants with a first acute or unspecified MI code and Fitbit and EHR data, 692 remained after excluding prior, post-MI or type 2 MI codes. After requiring at least 180 valid sleep nights in the year before the index date, the remaining exclusions and matching, the full matched cohort comprised 45 cases and 450 matched controls (10 per case; Table 1). Matching removed the pre-matching imbalance in age (standardised mean difference 0.91 before vs 0.05 after), diabetes (0.59 vs 0.00) and hypertension (0.86 vs 0.00) in the full matched cohort, in which BMI differed by −0.11 after matching (Table 1); in the subgroup with diabetes and/or hypertension, BMI differed by −0.36 and baseline sleep duration by +0.17; adjustment for these is reported below.

**Table 1.**
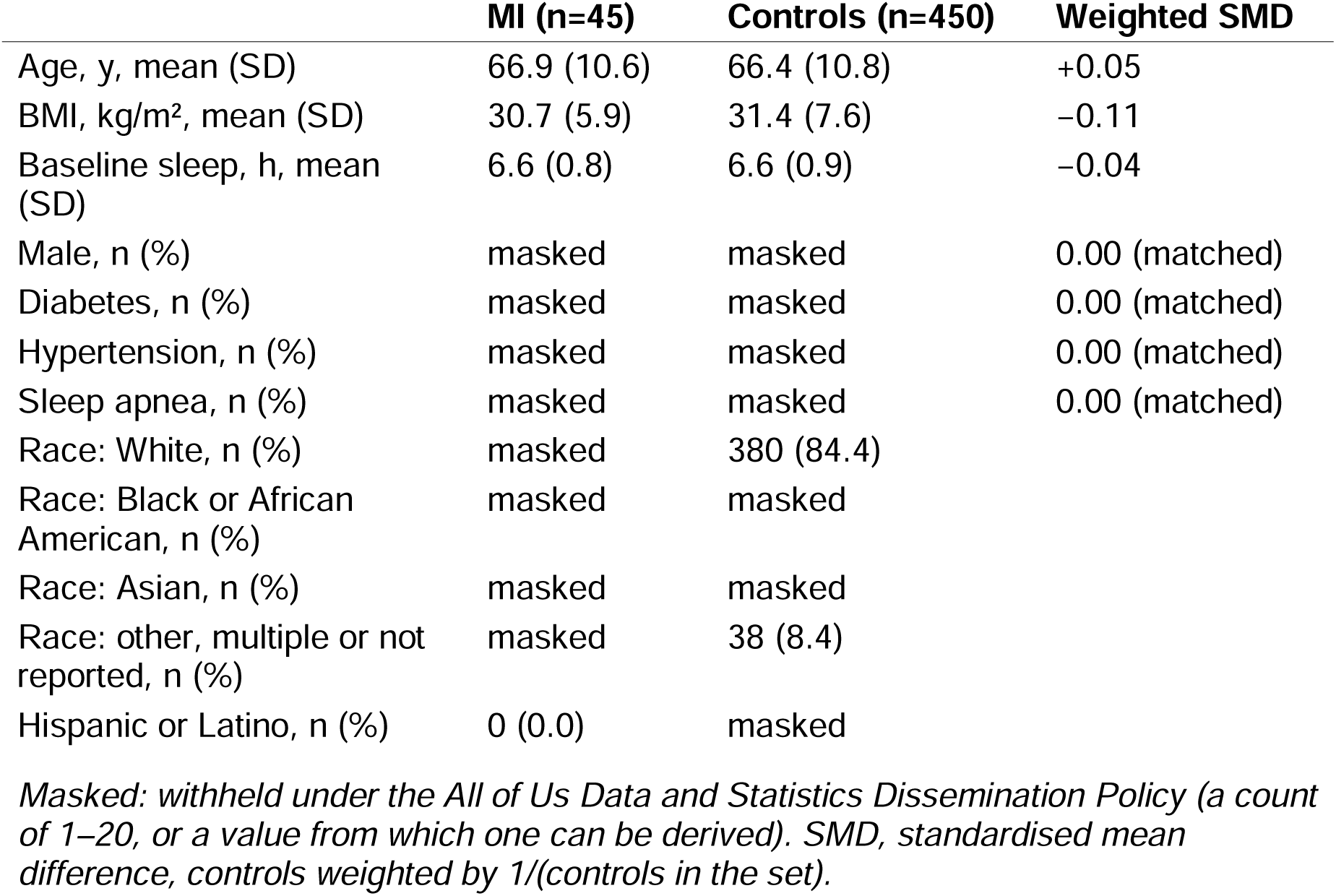
Characteristics of cases and matched controls (primary cohort)

Cases had more valid nights than controls over the year (median 290 [IQR 238–323] vs 244 [184–319]; δ 0.22, p=0.017), with a higher share of valid nights at baseline (78.2% vs 66.7%, p=0.012); the share of valid nights in the endpoint windows is given under Missing nights below.

### Main endpoints

REM-fraction variability changed transiently before MI, decreasing at days 30–16 and returning towards baseline over days 15–1; allowing for the choice of window, randomly placed index dates rarely reproduced this decrease-then-return pattern (max-T, p<0.001; Random index dates, below). At days 30–16, variability decreased in cases relative to their baseline and to controls (SD: δ −0.30, p=0.004; OR per SD 0.53, 95% CI 0.35– 0.80; RMSSD: δ −0.25, p=0.018; OR 0.55, 0.36–0.83), whereas changes in controls were small (|r| ≤ 0.08; Table 2, Figures 1b and 2). Before adjustment, the median SD of the nightly REM fraction in cases was 0.059 at baseline, 0.053 at days 30–16 and 0.062 at days 15–1, against 0.059, 0.060 and 0.059 in controls; for RMSSD, 0.080, 0.069 and 0.085 against 0.081, 0.081 and 0.082 (Table S1). From days 30–16 to days 15–1, variability increased in cases (RMSSD: δ +0.36, p<0.001; OR per SD 2.09, 1.32–3.31; SD: OR 1.90, 1.22–2.95; CV: OR 1.70, 1.13–2.55). Relative to baseline, variability at days 15–1 was higher in cases, but not significantly (endpoint 3: SD δ +0.14, p=0.17; RMSSD δ +0.17, p=0.10; CV δ +0.16, p=0.13; subgroup with diabetes and/or hypertension, δ +0.14 to +0.20, p≥0.086); the increase over the final 15 days thus largely reflects a return from the decrease at days 30–16. The decrease was not confined to cases with diabetes or hypertension: it was of the same size in the subgroup with either condition (SD δ −0.31, p=0.008) and in the same direction in sets without either condition (Table 2). Baseline-adjusted trajectories are shown in Figure 1c. Within-set permutation tests (1,000 permutations) gave similar results (SD: endpoint 1, p=0.003, endpoint 2, p=0.006; RMSSD: endpoint 1, p=0.006, endpoint 2, p=0.002; CV: endpoint 1, p=0.005, endpoint 2, p=0.014). Adjusting the conditional logistic regression for age, BMI and baseline sleep duration did not change the odds ratios (SD endpoint 1, 0.52, 0.34–0.79; RMSSD endpoint 2, 2.09, 1.32–3.33), nor did adjusting for nightly heart rate, wear and device (SD endpoint 1, 0.53, 0.34–0.83, against 0.54, 0.35–0.83, unadjusted in the same people; Table S2).

**Figure 2.**
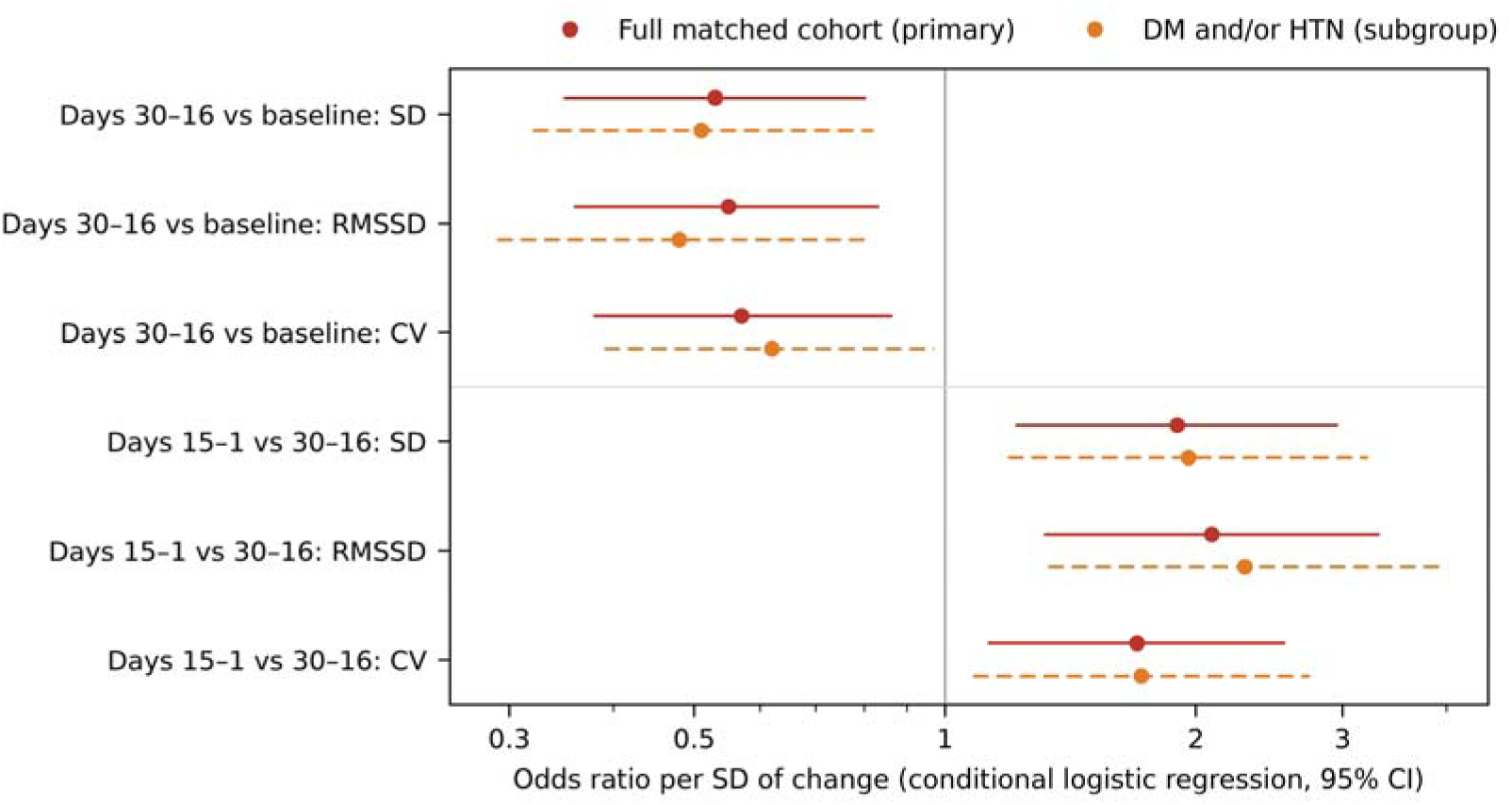
Odds ratio per SD of the change in REM-fraction variability, from conditional logistic regression within matched sets. Endpoint 1: days 30–16 vs baseline; endpoint 2: days 15–1 vs days 30–16.

**Table 2.**
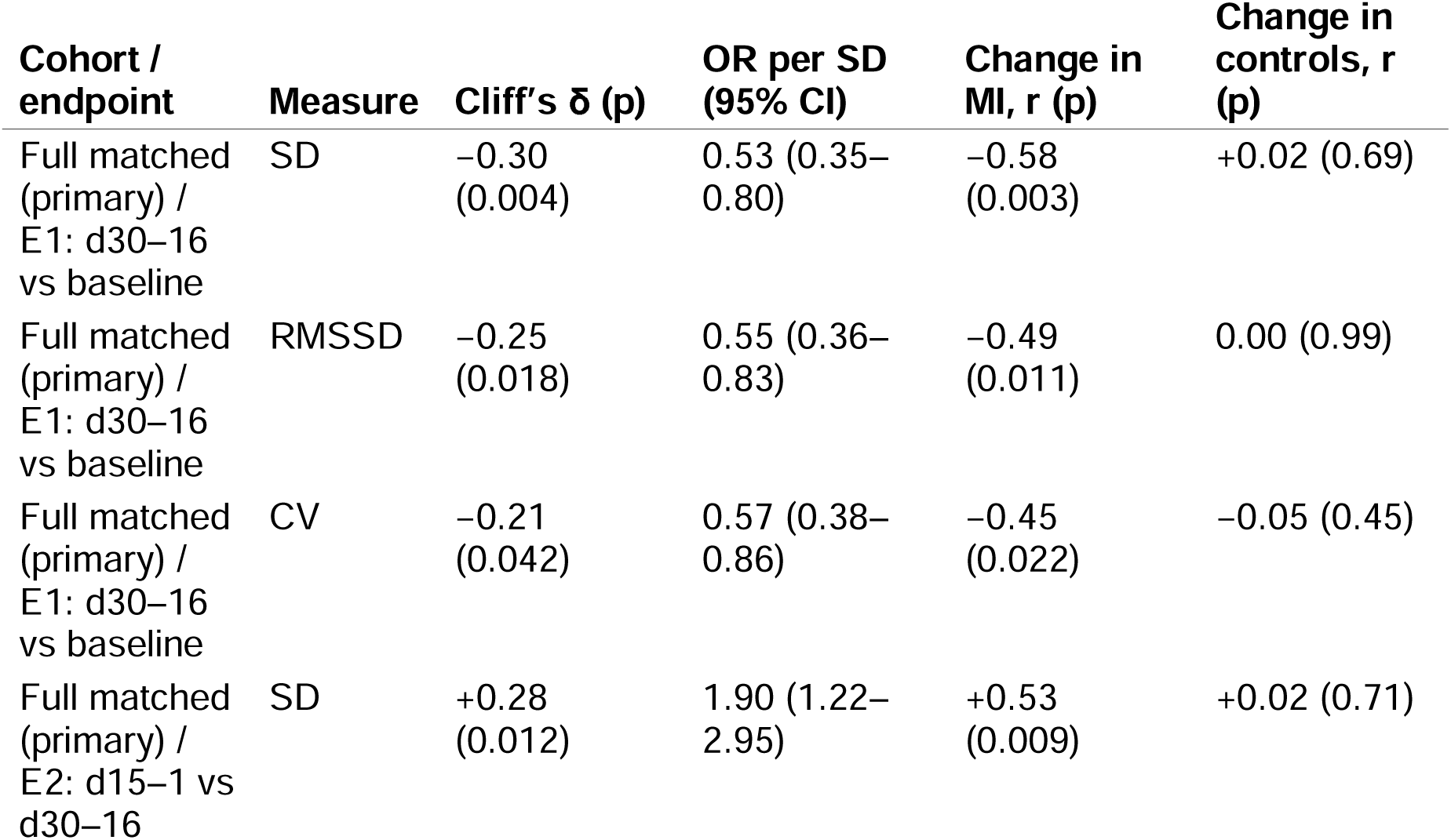
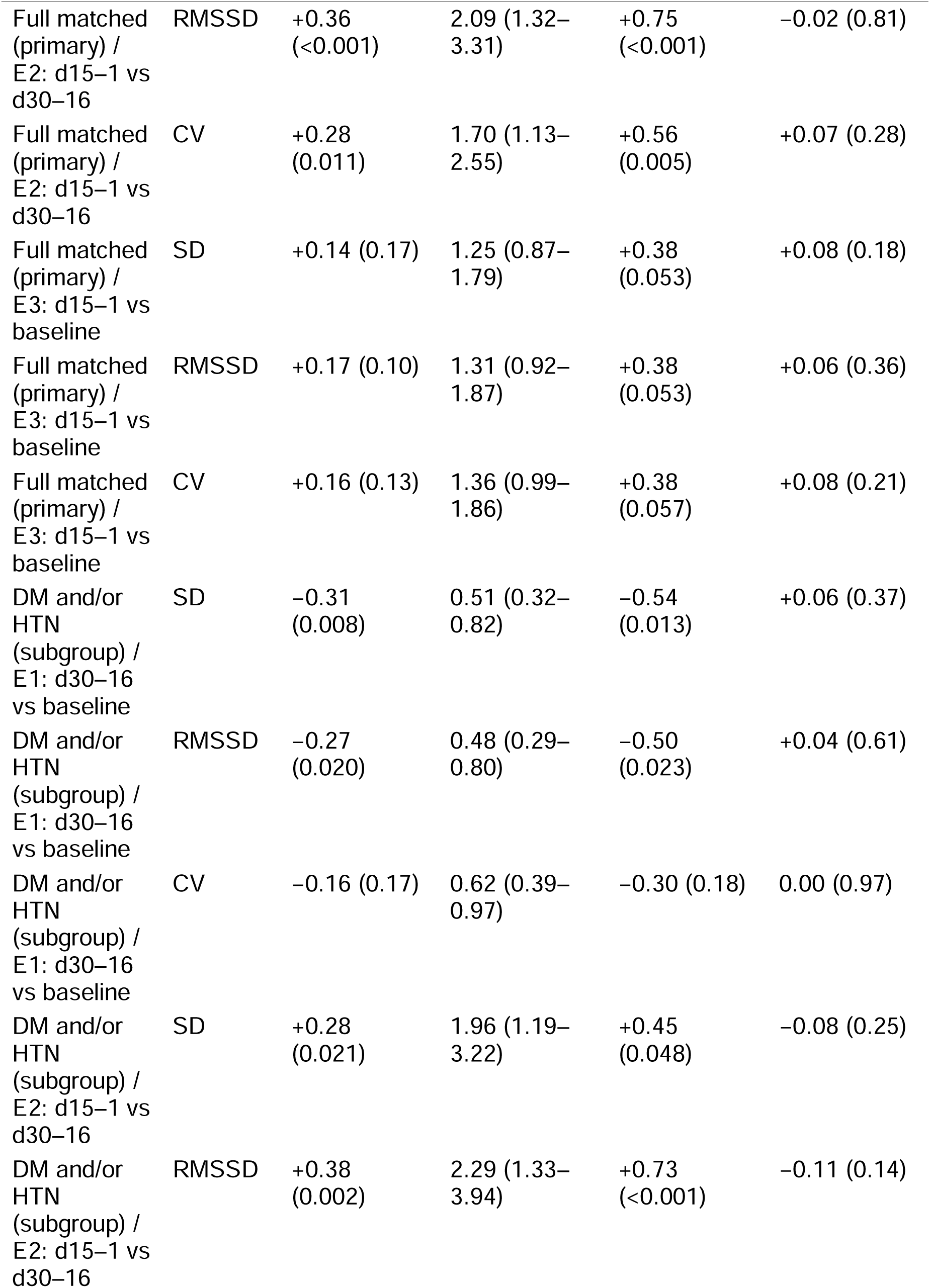
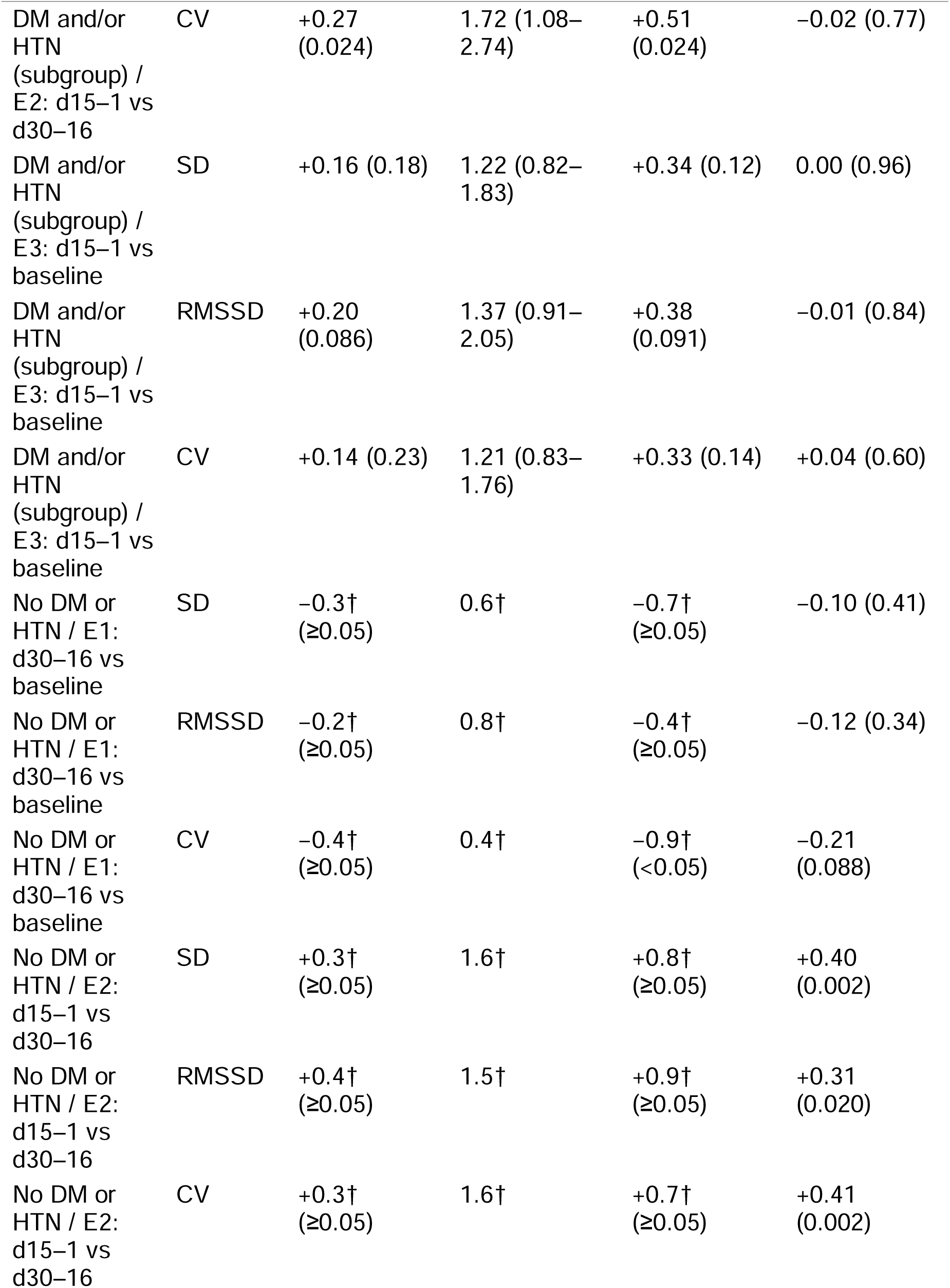
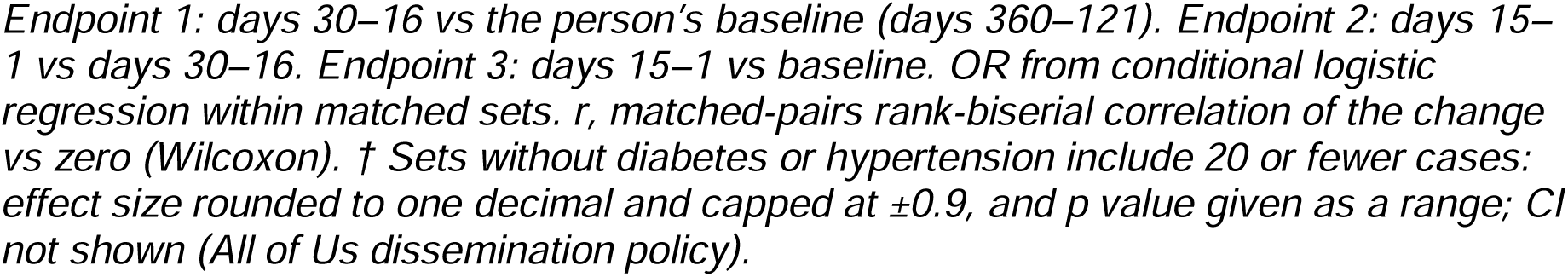
Main endpoints: change in REM-fraction variability, cases vs matched controls.

### Per-person detection

In the full matched cohort, per-person z scores discriminated cases from controls with AUCs of 0.51–0.66; the best was RMSSD with either a drop at days 30–16 or a rise at days 15–1 (AUC 0.66; OR for being flagged 3.44, 1.34–8.85). In the subgroup with diabetes and/or hypertension the same rule reached an AUC of 0.64 (OR 2.28, 0.75– 6.92). With continuous monitoring over days 120–1, controls had about 2.3 false alarms per year (2.2 in the subgroup). Sensitivity and the false-positive rate could not be reported because, together with the odds ratio, they would reveal counts of 20 or fewer (Table 3).

**Table 3.**
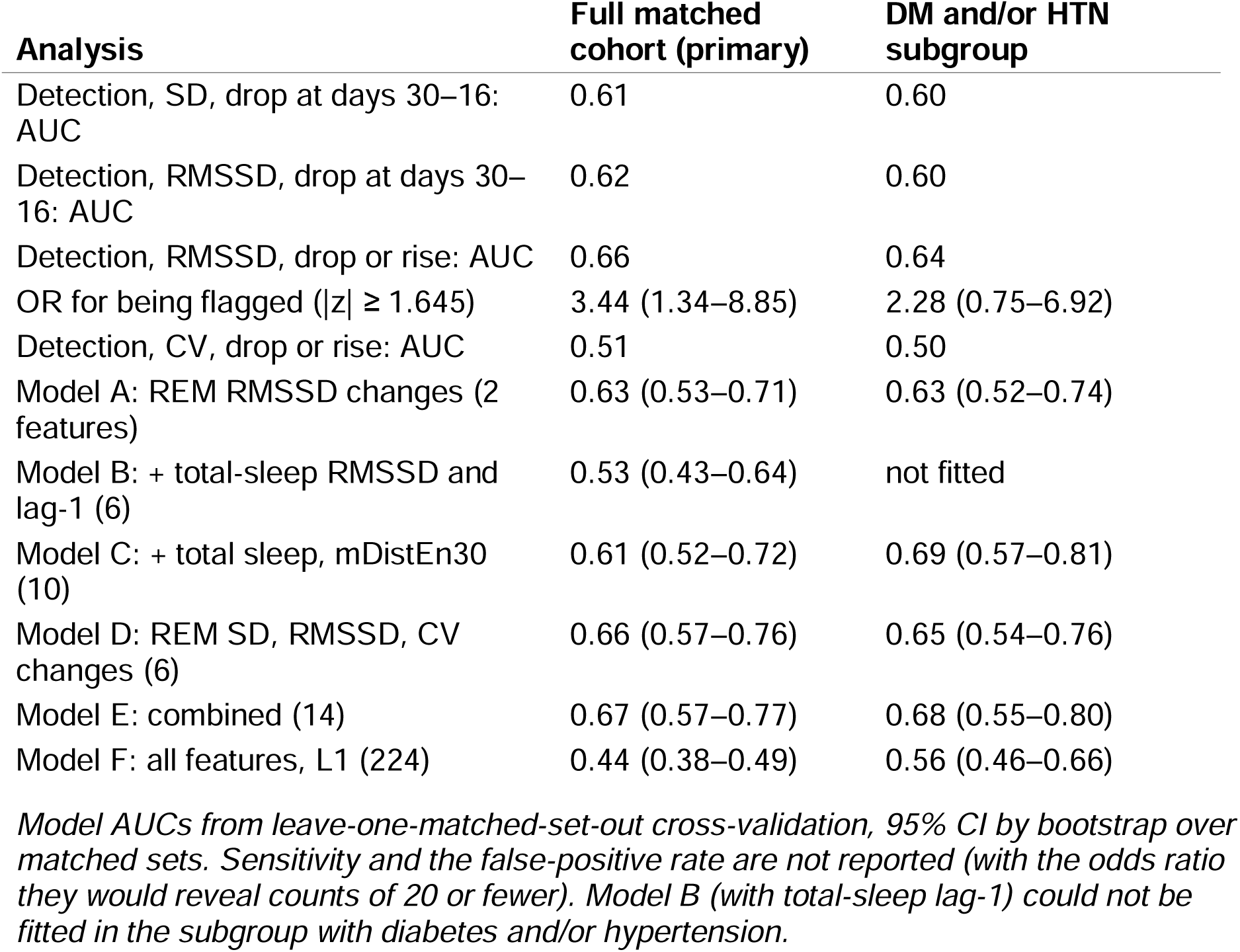
Per-person detection and cross-validated person-level models.

The dip alone separated MI cases not only from their own controls but also from the cases of other acute events (Table S3). In the full matched cohort, the dip in SD discriminated MI cases from their own controls with an AUC of 0.61 (95% CI 0.51–0.71) and from the cases of pneumonia, fracture or appendicitis and any admission with AUCs of 0.65, 0.65 and 0.66; the dip in RMSSD gave 0.62 (0.51–0.73) against own controls and 0.71 (0.60–0.81) against fracture or appendicitis. Composite indices that also required a subsequent rise performed similarly (SD: 0.62, 0.51–0.73, against own controls).

### Other acute events

The decrease in SD at days 30–16 was not seen at the same time before the other acute events examined (Table S4, Figures 1b and 1d). In the full matched cohort, endpoint 1 for SD was δ −0.30 (95% CI −0.47 to −0.12) before MI, against −0.05 (−0.22 to +0.11) before pneumonia admission (69 cases, 690 controls), +0.04 (−0.07 to +0.15) before fracture or appendicitis admission (130 cases, 1,298 controls) and +0.02 (−0.03 to +0.06) before admission of any cause (859 cases, 8,247 controls); pneumonia and any admission excluded admissions with an acute MI code. Compared directly, the change before MI was larger than before each (difference in δ −0.25, −0.49 to −0.01; −0.34, −0.55 to −0.13; −0.32, −0.49 to −0.14). RMSSD also tended to decrease before pneumonia, by less than before MI (δ −0.17, −0.31 to −0.01; MI δ −0.25; difference −0.08, −0.33 to +0.18), so RMSSD was not used as evidence of specificity. The increase over days 15–1 was shared with pneumonia (SD endpoint 2: δ +0.26, p=0.005; RMSSD: δ +0.27, p=0.005) but not with fracture or appendicitis or any admission (Table S5). Before ischaemic stroke (22 cases, 220 controls) and heart-failure admission (27 cases, 270 controls), endpoint 1 estimates were close to zero or positive (δ about 0 to +0.1), but power to detect an effect of the size seen in MI was at most 0.64 and 0.73. Results were similar in the subgroup with diabetes and/or hypertension (pneumonia, SD endpoint 1: δ −0.07, −0.26 to +0.11).

### Cases without recorded symptoms or treatment

In the full matched cohort, SD decreased at days 30–16 in matched sets whose case had no angina or chest-pain code and no newly started cardiovascular drug in the 90 days before the index date (δ −0.30, p=0.011), and variability increased over days 15–1 (SD: δ +0.24, p=0.050; RMSSD: δ +0.34, p=0.006) (Table S6). In the subgroup with diabetes and/or hypertension, 20 or fewer cases had no recorded symptoms or treatment; in these matched sets, REM-fraction SD and RMSSD decreased at days 30– 16 (δ about −0.3, p<0.05). The changes were therefore of similar size in cases without recorded symptoms or treatment started before the event.

### Heart failure

When cases and controls with a heart-failure code were included (heart failure neither excluded nor matched on), the main endpoints were unchanged (full matched cohort: SD endpoint 1, δ −0.29, p=0.004; RMSSD endpoint 2, δ +0.35, p<0.001; Table S7), and adding heart-failure status as a covariate did not change the odds ratios (SD endpoint 1, 0.51 unadjusted and 0.51 adjusted; RMSSD endpoint 2, 2.04 and 2.05). In that cohort, controls with a heart-failure code did not differ significantly from those without in any endpoint change (p≥0.05). For the per-person SD-drop score, MI cases were separated from controls without heart failure (AUC 0.64) but not from controls with heart failure (AUC 0.5, rounded to one decimal because 20 or fewer controls with heart failure had a value).

### Exploratory analyses

In leave-one-set-out cross-validation in the full matched cohort, a model with REM-fraction RMSSD changes alone (model A) reached an AUC of 0.63 (0.53–0.71); adding total-sleep variability and lag-1 autocorrelation (model B) gave 0.53 (0.43–0.64), and further adding 30-night distribution entropy of the light and REM fractions (model C) 0.61 (0.52–0.72). The SD, RMSSD and CV of the REM fraction (model D) gave 0.66 (0.57– 0.76), and model E, combining models C and D, the highest AUC, 0.67 (0.57–0.77). An L1-penalised model including all 224 features did not discriminate (AUC 0.44, 0.38– 0.49). In the subgroup with diabetes and/or hypertension, model C gave the highest AUC (0.69, 0.57–0.81) (Table 3).

In the exploratory segment analysis of the full matched cohort (756 tests, 44 with p<0.05, about 38 expected by chance), the mean REM fraction did not change (days 30–16: δ −0.08, p=0.46), so the change was in variability rather than level. The wake fraction of time in bed changed before the event: its minimum fell over days 60–31 (δ −0.27, p=0.008), its lag-1 autocorrelation tended to rise at days 30–16 (δ +0.17, p=0.094) and its minimum rose again over days 30–1 relative to days 60–31 (δ +0.37, p=0.001). Results were similar in the subgroup with diabetes and/or hypertension (39 of 756 tests with p<0.05; minimum of the wake fraction over days 60–31, δ −0.30, p=0.011).

Among complexity measures, 2 of 80 contrasts had p<0.05 in the full matched cohort (about 4 expected by chance): 30-night bubble entropy of total sleep increased at days 30–16 (δ +0.22, p=0.044), and 30-night distribution entropy of the light-sleep fraction fell over days 15–1 (δ −0.25, p=0.041) after a non-significant rise at days 30–16 (δ +0.15, p=0.17). In the subgroup with diabetes and/or hypertension, 3 of 68 contrasts had p<0.05 (about 3 expected), among them an increase in 30-night distribution entropy of the light-sleep fraction at days 30–16 (δ +0.28, p=0.029; OR per SD 1.46) and in the changepoint probability of the light-sleep fraction over days 15–1 (δ +0.26, p=0.013). Lag-1 autocorrelation of total sleep over the 21 nights before the index date did not differ (full matched cohort, δ −0.09, p=0.50). Day-level classifiers predicting the event 1–14 days ahead did not discriminate (AUC 0.44–0.56).

### Robustness checks

#### Random index dates

Randomly placed index dates did not reproduce the observed pattern. In the full matched cohort, endpoint 1 for SD was δ −0.28 against a mean of +0.03 across random dates (central 95%, −0.14 to +0.20; p=0.0012). Allowing for the choice of window among the eight possible positions, the decrease-then-increase in SD (max-T, p<0.001), the decrease alone (SD, p=0.0034) and the increase in RMSSD (p=0.0035) remained significant (observed values refer to the matched sets with at least one usable random date). Max-T standardises each window by the mean and SD of its own random-date distribution (Table S8); without standardisation, the decrease alone gave p=0.047 (0.11 in the subgroup with diabetes and/or hypertension), because the random-date means differed between windows (−0.08 to +0.04) while their SDs were similar (0.088–0.096). One-sided, max-T gave p=0.0019 for the decrease alone, p<0.001 for the decrease-then-increase and p=0.0019 for the increase in RMSSD; over the three windows of the range suggested by the earlier data release (days 45–31, 30– 16 and 15–1), it gave p=0.0013, p<0.001 and p=0.0016. Day by day, the decrease was not detected as a continuous change: in a cluster permutation test of the 14-night rolling SD over days 120–1, the larger of the two clusters of days with p<0.05 within days 30– 16 lasted 2 days (days 17–16; mass 0.21; p=0.58), against a median largest cluster of 3 days (95th percentile 13) at random dates. One-sided, the decrease cluster (days 20– 15) had p=0.41, and with a forming threshold of p<0.01 no cluster formed. Adjacent days of a 14-night rolling window are strongly correlated, which lengthens clusters arising by chance, and the change itself lasted less than two weeks, so this test had little power. In random halves of the matched sets, the window with the largest change in one half was days 30–16 in 776 of 1,000 splits, and the change was preserved in the other half (median δ −0.25, 95% interval −0.41 to +0.23). The decrease in RMSSD was attenuated in the held-out halves (median δ −0.08, 95% interval −0.28 to +0.15). With the shorter baseline, the decrease in SD was also rarely matched at random dates (single window, p<0.001).

#### Consistency across endpoints

Restricted to persons with all three endpoints, endpoints 1 and 2 were similar (SD endpoint 1, δ −0.30, p=0.005; RMSSD endpoint 2, δ +0.36, p<0.001).

#### Index date

The first MI code was usually recorded on the day the visit began; where the visit had begun 1–3 days earlier with an MI presentation, moving the index date to the visit start did not change the results (SD endpoint 1, δ −0.30, p=0.004; RMSSD endpoint 2, δ +0.37, p<0.001). Moving all windows 3 days earlier left both endpoints largely unchanged (SD endpoint 1, δ −0.22, p=0.032; RMSSD endpoint 2, δ +0.35, p=0.002). Moving them 5 days earlier attenuated both endpoints (SD endpoint 1, δ −0.16, p=0.12, partly because the window then covers days 35–21; RMSSD endpoint 2, δ +0.19, p=0.083).

#### Index at the first angina code

In matched sets whose case had a first angina code in the 90 days before the MI, the whole set was re-indexed to that code. The results were unchanged (full matched cohort: SD endpoint 1, δ −0.31, p=0.003; RMSSD endpoint 2, δ +0.39, p<0.001; original index, δ −0.30 and +0.36), as they were with the first angina or chest-pain code and with these sets removed (SD endpoint 1: δ −0.32, p=0.003) (Table S9). Too few cases were re-indexed to tell whether the timing of the changes follows the angina code or the MI.

#### Missing nights

At days 30–16, the share of valid nights did not differ between cases and controls (73.3% vs 77.3%; δ −0.07, p=0.46), so the decrease in variability is not explained by selective data loss. At days 15–1, cases had somewhat fewer valid nights (67.6% vs 75.4%; δ −0.15, p=0.086), with more nights without a main-sleep record (23.6% vs 17.9%; δ +0.13, p=0.13), but neither difference was significant. Nights with zero REM sleep made up under 1% of nights in every window and are included as a REM fraction of 0; RMSSD uses only pairs of consecutive calendar nights.

## Discussion

In this matched case–control study using long-term Fitbit data in the All of Us Research Program, night-to-night variability of the REM-sleep fraction changed within individuals in the weeks before MI: it decreased relative to their own baseline at days 30–16 and returned towards baseline over the final 15 days, while changes in matched controls were small. Scored against each person’s own baseline, these changes discriminated cases from controls with AUCs of about 0.6–0.7, although these are probably optimistic because the best rule used RMSSD, whose decrease was attenuated in held-out data.

The decrease in REM-fraction SD was not seen at the same time (days 30–16) before admissions for pneumonia, fracture or appendicitis or any cause, it was larger before MI than before each of them and it persisted in cases without recorded symptoms or treatment and when the index date was moved to the first angina code, which argues against a non-specific effect of acute illness, hospital admission, contact with health care or treatment. The increase over the final two weeks, in contrast, was also seen before pneumonia and probably reflects acute illness more generally. RMSSD also decreased before pneumonia, and its difference from MI was not significant, so only the SD of the REM fraction supports specificity. Comparisons with ischaemic stroke and heart-failure admission were too small to be informative. When people with heart failure were included in the MI analysis, the changes were unchanged, and MI cases could not be distinguished from controls with heart failure. The decrease was of the same size in the subgroup with diabetes and/or hypertension and in the same direction in the sets whose case had neither condition, so it may be a feature of MI itself rather than of the comorbid state.

Variability, rather than the mean level, has been the signal in every analysis of these data: between groups in the earlier data release, and within persons here. What changed before MI is therefore best described as the night-to-night consistency of wearable-estimated sleep architecture rather than its average composition. The difference between groups in the earlier release was later found to reflect diabetes and hypertension; the evidence that the change is linked to the MI itself comes from the within-person analysis, whose decrease-then-return pattern held against randomly placed index dates and was absent before other acute events. Reduced variability has been linked to cardiovascular risk on other timescales: reduced heart-rate variability predicts mortality after MI ^25^ and cardiac events in the general population ^26^, and a loss of variability and complexity of physiological signals, measured for example by entropy, has been proposed as a general sign of failing regulation ^27^. These measures concern beat-to-beat fluctuations over seconds to minutes, whereas the change here was in the night-to-night variation of a wearable-estimated sleep stage, so the parallel is one of form rather than of mechanism. Because Fitbit estimates sleep stages partly from heart rate and movement, a change in nocturnal heart-rate dynamics could contribute to the staging output; adjusting for nightly heart rate (its mean and night-to-night variability), wear and device did not change the result, and further heart-rate analyses are the subject of ongoing work.

In the same data source, the percentage of REM sleep fell before incident heart failure ^14^. Before MI, in contrast, the mean REM fraction did not change, whereas its night-to-night variability did.

These changes match the prodromal literature in timing: fatigue and sleep disturbance are reported weeks before MI ^5,6^, and we found within-person changes in sleep architecture over a similar window, 2–4 weeks before the event, in cases with and without recorded symptoms. They were detectable with simple, surface-level measures — the SD and RMSSD of the REM fraction — whereas more complex dynamical measures and day-level classifiers added little. Wearables thus complement long-term risk profiling from habitual sleep ^8^ with short-term, within-person change.

REM sleep is accompanied by surges of sympathetic activity ^28^ that may contribute to nocturnal cardiovascular events, so changes in REM regulation are plausibly linked to cardiovascular instability. One interpretation of the decrease at days 30–16 is a period of steadier sleep preceding the event; in the exploratory analyses the wake fraction of time in bed also tended to become more persistent from night to night over the same window (δ +0.17, p=0.094), although this was one of many exploratory contrasts. Plaque instability frequently precedes occlusive coronary thrombosis by days to weeks ^29^, but how an unstable plaque could be followed by such a sequence is not known, and the following are hypotheses only. Our main hypothesis is that inflammation in the plaque and the circulation shifts autonomic balance, including through the vagal anti-inflammatory reflex, and that cytokines act on sleep regulation; an alternative is that silent ischaemia or microembolism during plaque instability alters afferent input from the heart to brainstem networks that govern arousal and transitions between REM and non-REM sleep. Both need direct testing.

The change was detectable at the level of the individual with simple measures scored against the person’s own baseline (AUCs of about 0.6–0.7 from a single feature). These AUCs are probably optimistic: the best rule used RMSSD, whose decrease was attenuated in held-out halves of the matched sets, whereas the decrease in SD was preserved. Whether a person-specific rule of this kind can contribute to wearable-based early warning needs larger cohorts and prospective studies.

## Limitations

The full matched cohort was small (45 cases). Requiring at least 180 valid nights in the year before MI excluded most of the 692 eligible cases, so participants with sparse wearable use are not represented, and cases wore the device more consistently than controls at baseline (78.2% vs 66.7% valid nights), which may be related to health status. MI and comorbidities were identified from EHR codes; sleep apnea and heart failure are likely under-recorded, and the index date is the date of the first code rather than symptom onset, although moving it to the start of the acute visit or to the first angina code did not change the results. Fitbit sleep staging agrees only moderately with polysomnography ^10^, the proprietary staging algorithm may have changed over the study period and, because staging uses heart rate and movement, our findings concern wearable-estimated rather than polysomnographic REM sleep. Cases had somewhat fewer valid nights than controls in the final 15 days, although not significantly; windows were compared with equal numbers of nights, but data missing before the event may not be missing at random. The endpoint windows were chosen after an exploratory analysis of these data, refining a decrease seen in an earlier, overlapping data release, although randomly placed index dates did not reproduce the finding even after allowing for the choice of window; the timing of days 30–16 is therefore exploratory and is to be confirmed, with the window fixed in advance, in new cases of a later All of Us data release. Secondary and exploratory analyses were not adjusted for multiple testing. There was no external validation cohort, and Apple Watch data in All of Us included too few MI cases with sleep data for validation. The results come from All of Us participants who shared Fitbit data and wore the device on most nights; they were older, mostly

White and often obese, and other devices and populations were not studied. Under the All of Us dissemination policy, sensitivity and several descriptive characteristics could not be reported. A small number of cases had a separate acute illness recorded before the MI code and may have had the MI during a hospital stay. Comparisons with ischaemic stroke and heart-failure admission were underpowered.

## Conclusions

In adults who use consumer wearables, a first MI was preceded by a transient change in night-to-night REM-sleep variability, a decrease at days 30–16 and a return over the following two weeks. The decrease was not seen at the same time before other acute admissions, did not depend on recorded symptoms or treatment, did not appear to be confined to people with diabetes or hypertension and was not explained by nightly heart rate, wear or device. This transient change can be detected against each person’s own baseline; its timing, and whether it can support wearable-based early warning, are to be confirmed in new cases.

## Declarations

## Supporting information

Graphical abstract

## Acknowledgements

We gratefully acknowledge All of Us participants for their contributions, without whom this research would not have been possible. We also thank the National Institutes of Health’s All of Us Research Program for making available the participant data examined in this study.

## Data availability

Data are available to authorised researchers through the All of Us Researcher Workbench (Registered Tier Dataset v9). Participant-level data cannot be shared by the authors.

## Code availability

The analysis code is available from the corresponding authors on request.

## Ethics

This study was approved by the Medical Research Ethics Committee of the Institute of Science Tokyo as part of the protocol “Prediction of heart failure and multimorbidity states using biobank samples and information, and epigenomic analysis of haematopoietic stem cells and monocytes” (I2026-142). All of Us data are de-identified and were accessed under the All of Us Data User Code of Conduct; no additional informed consent was required.

## Author contributions

E.I. and K. Fujiu designed the study; E.I. performed the analyses; J.S., K.U., N.S., H.S., R.U., Y.Y., Z.L. and S.M. contributed to the data analysis and manuscript preparation; E.I. and K. Fujiu interpreted the results and wrote the manuscript; K. Fujiwara and K.A. interpreted the results and revised the manuscript.

## Supplementary material

**Figure S1.**
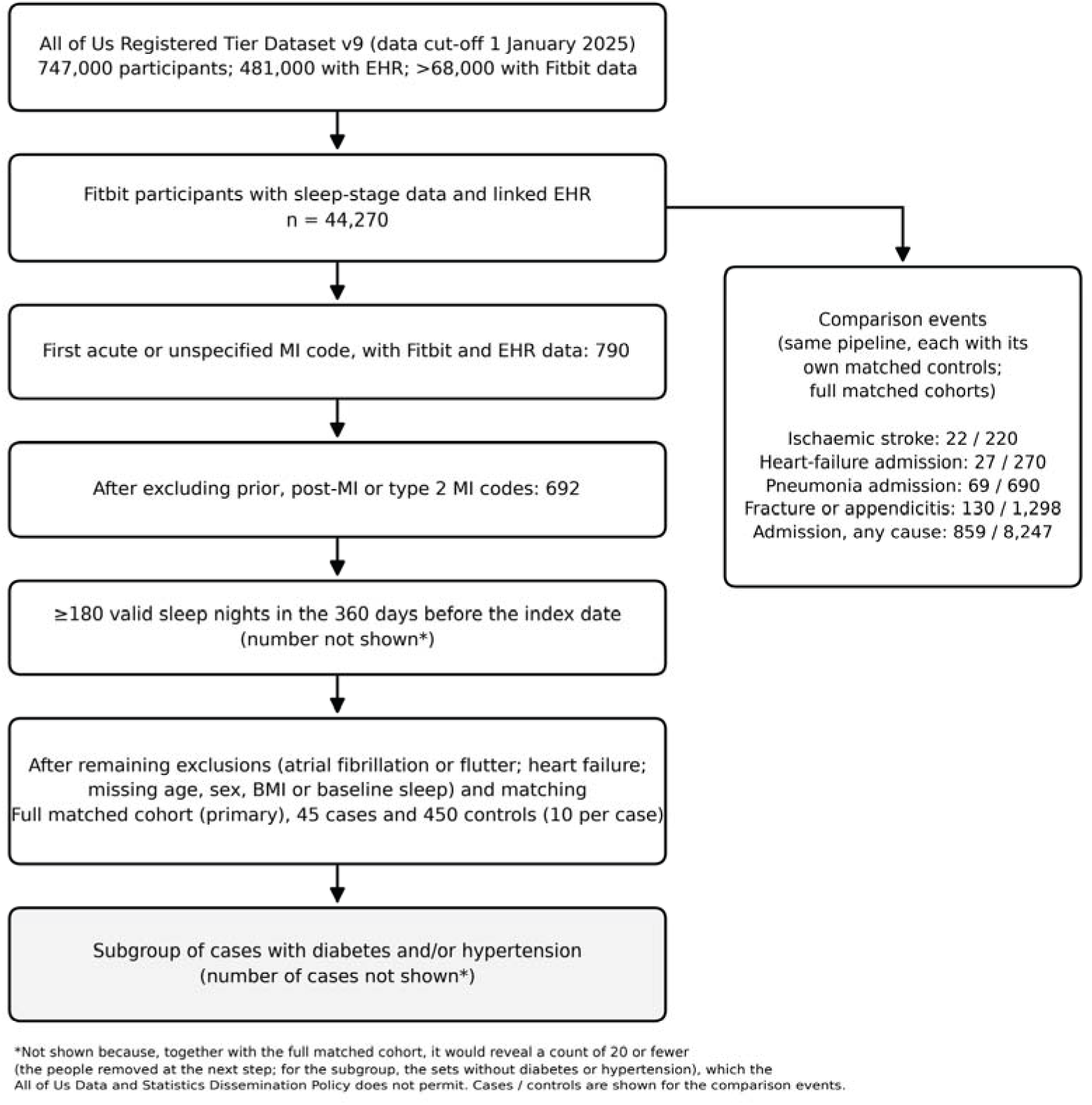
Participant flow. Numbers of participants at each step from All of Us Registered Tier Dataset v9 to the full matched cohort (primary), and the comparison event groups analysed with the same pipeline.

---

**Table S1.**
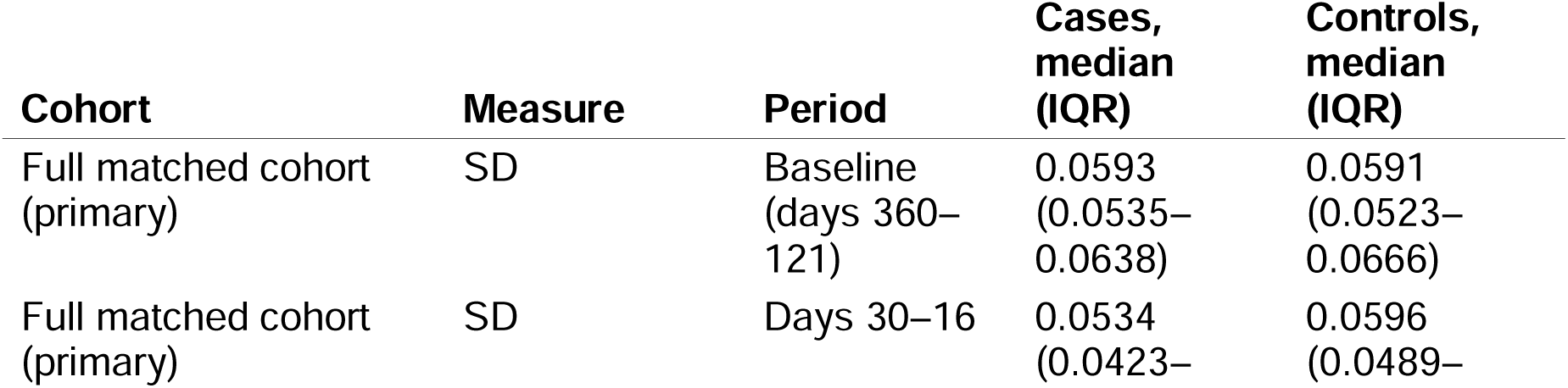
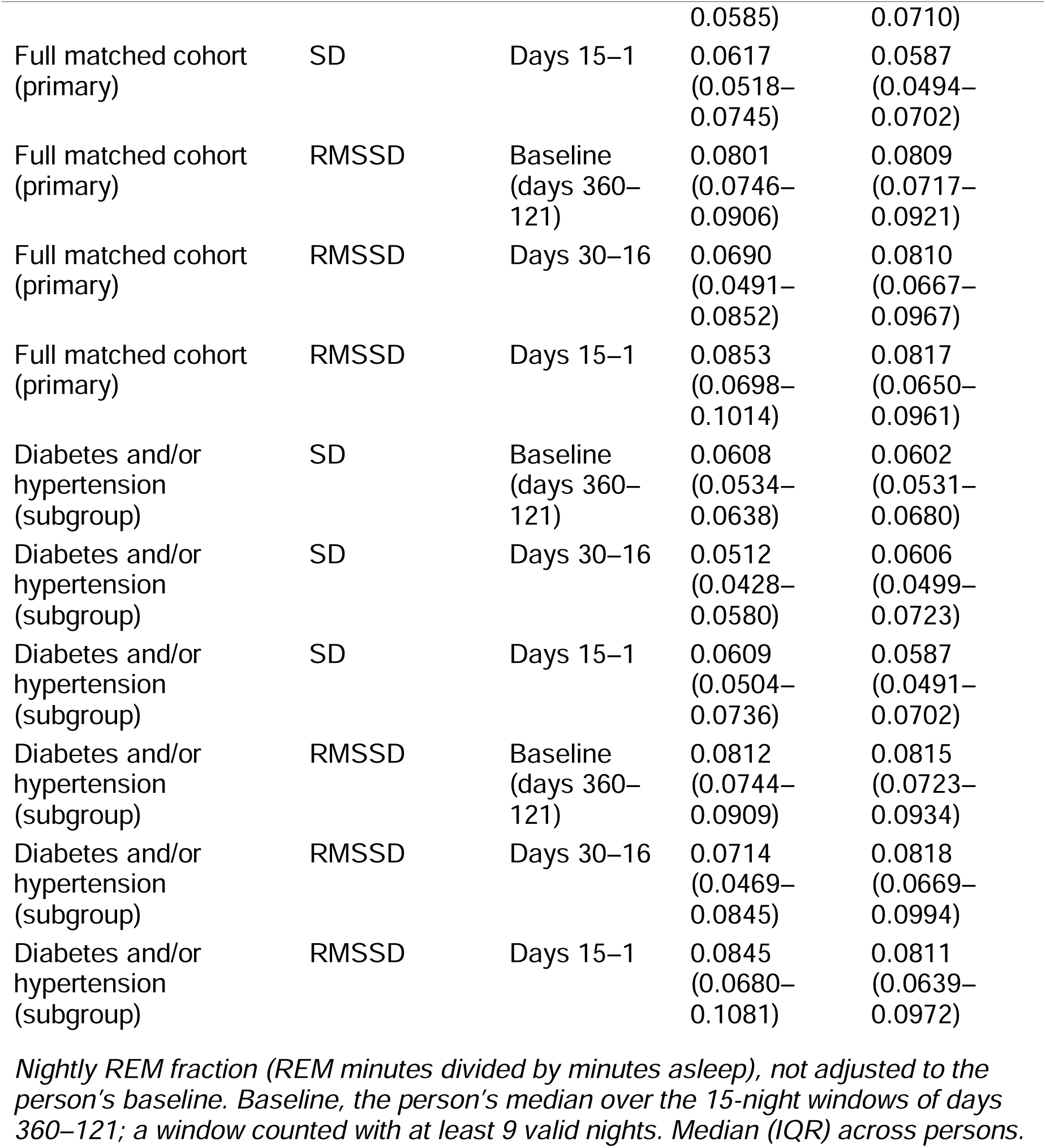
REM-fraction SD and RMSSD before adjustment, by period.

**Table S2.**
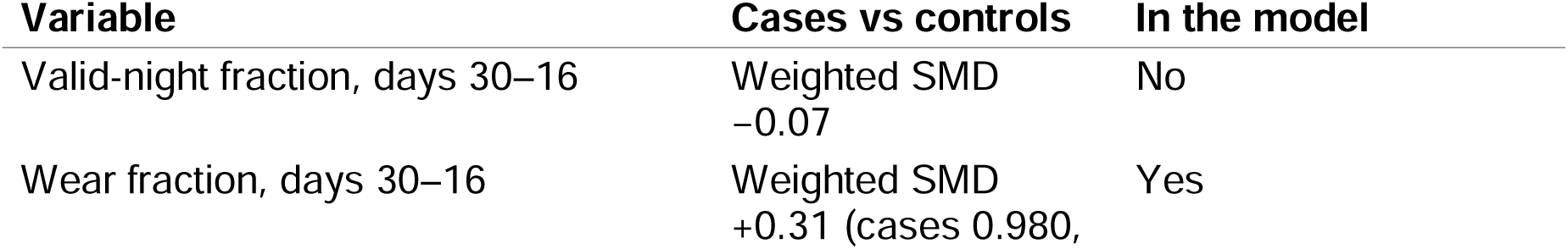
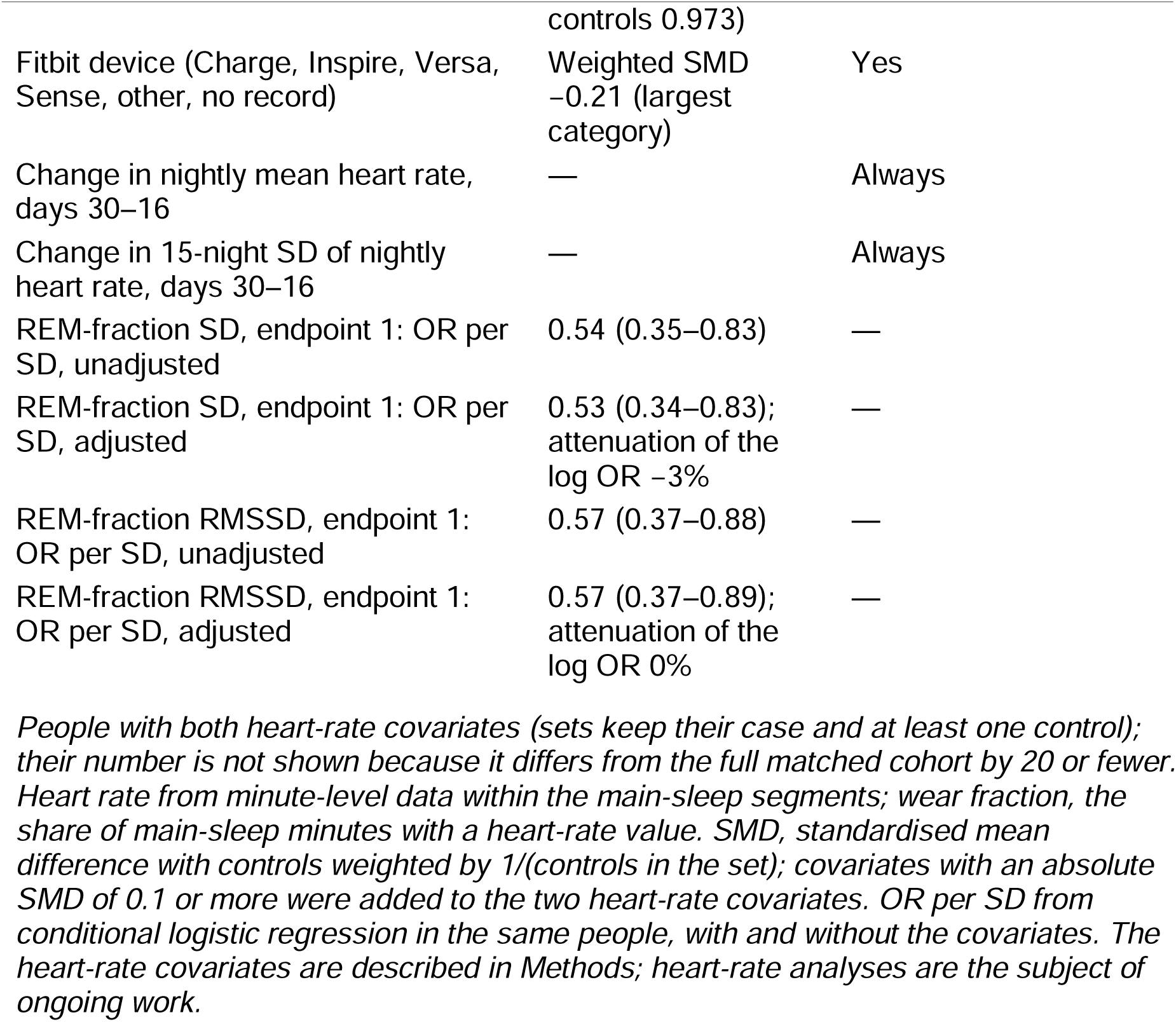
Adjustment for nightly heart rate, wear and device (full matched cohort)

**Table S3.**
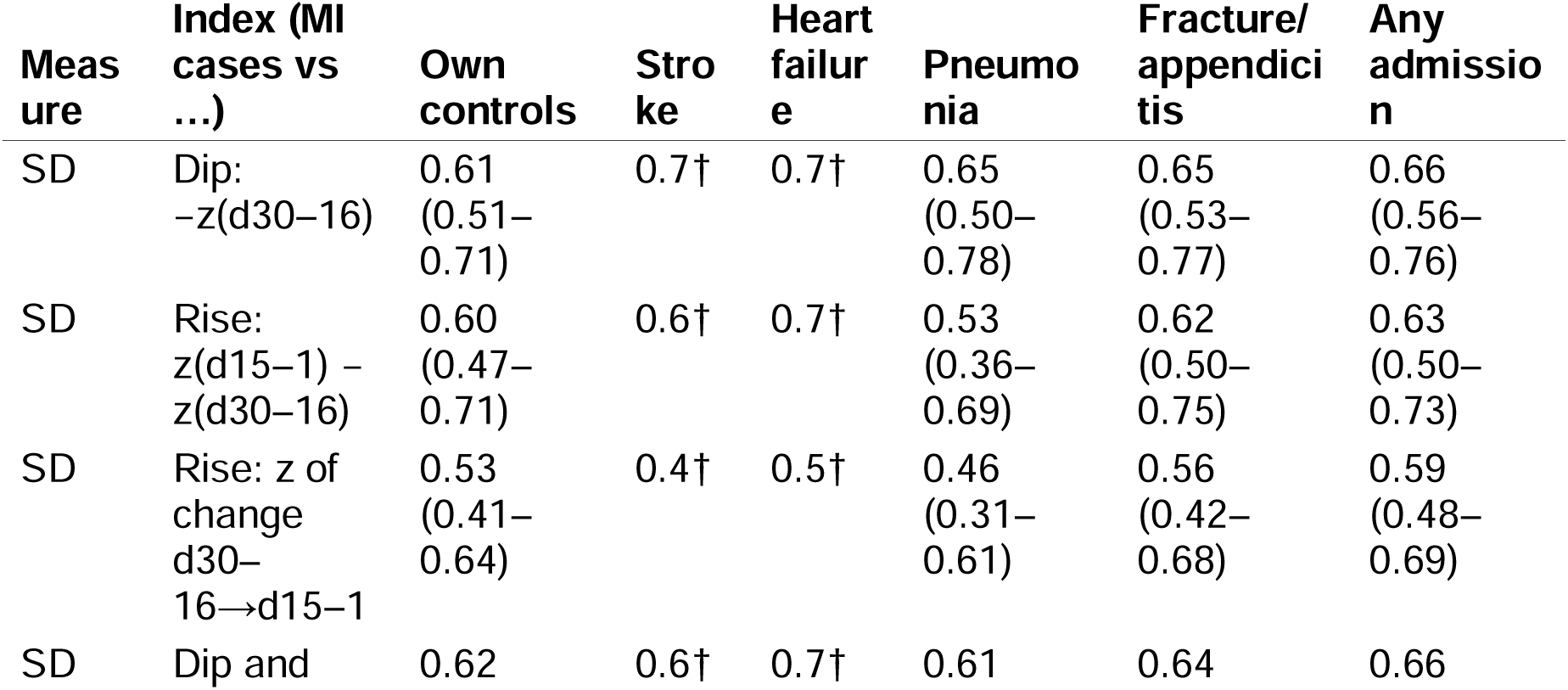
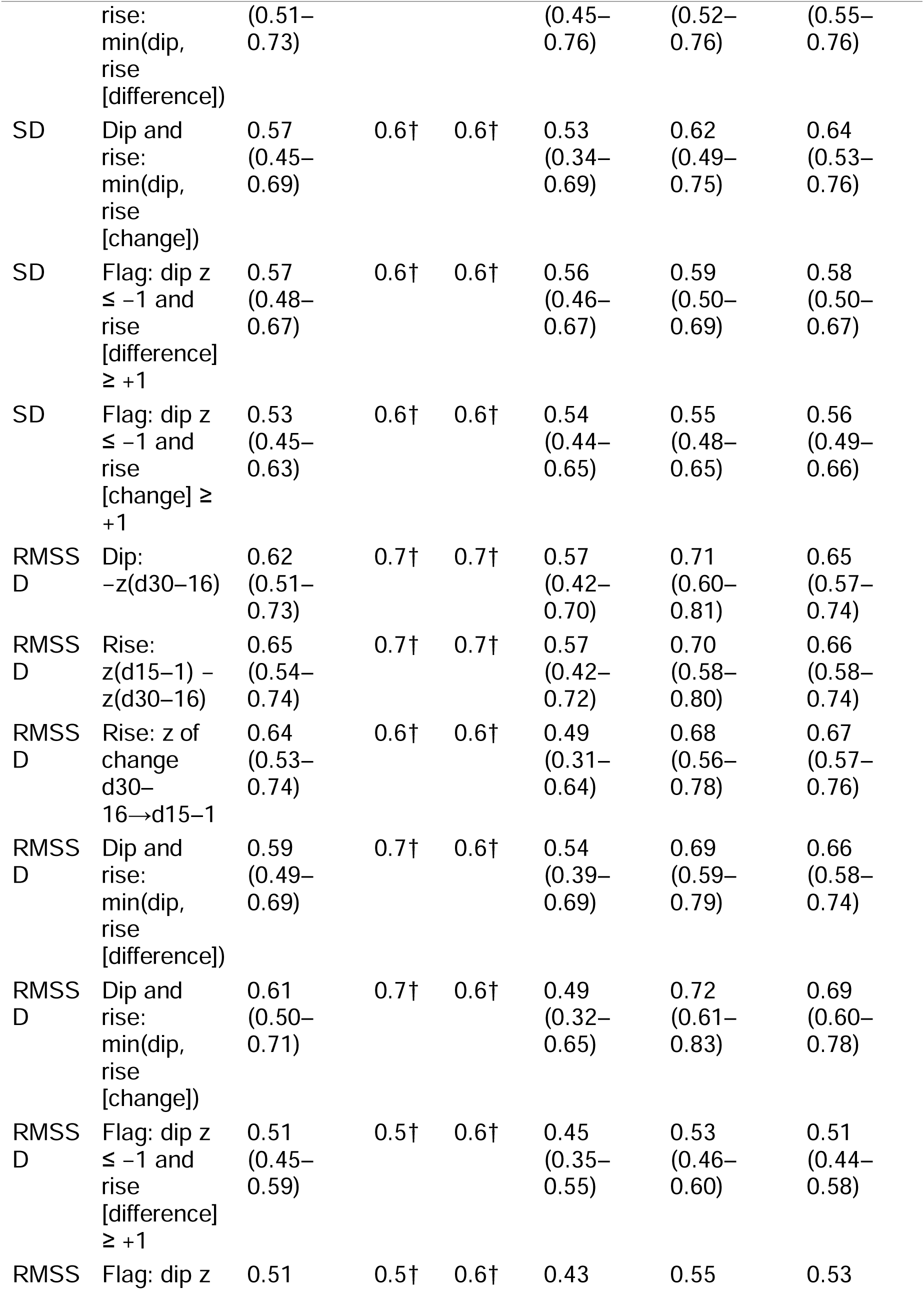
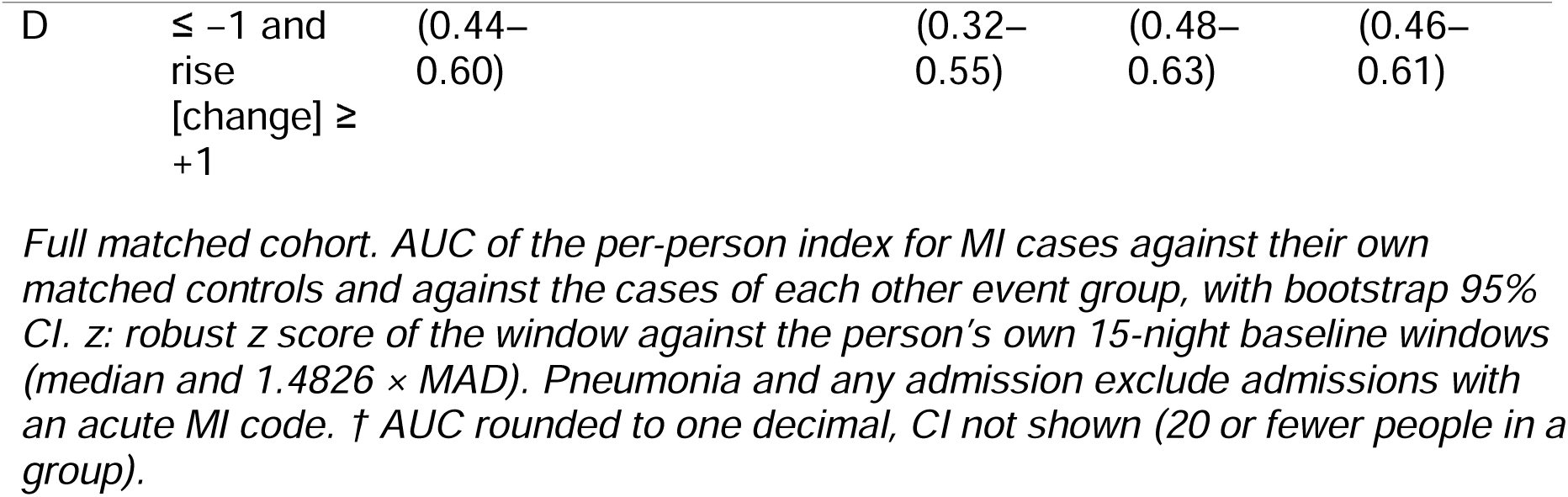
Composite per-person indices: AUC for MI cases vs own controls and other events.

**Table S4.** REM-fraction variability before MI and other acute events (full matched cohort)

| Event | Cases / controls | SD, E1: $\delta$ (95% CI) | RMSSD, E1: $\delta$ (95% CI) | SD, E2: $\delta$ (p) | RMSSD, E2: $\delta$ (p) | Power, $\delta -0.30$ |
| --- | --- | --- | --- | --- | --- | --- |
| MI | 45 / 450 | –0.30<br>(–0.47 to –0.12) | –0.25<br>(–0.42 to –0.05) | +0.28<br>(0.012) | +0.36<br>($<0.001$) | — |
| Ischaemic stroke | 22 / 220 | +0.1†<br>($\geq 0.05$) | +0.1†<br>($\geq 0.05$) | +0.1†<br>($\geq 0.05$) | 0.0†<br>($\geq 0.05$) | $\leq 0.64$ |
| Heart-failure admission | 27 / 270 | 0.0†<br>($\geq 0.05$) | 0.0†<br>($\geq 0.05$) | +0.2†<br>($\geq 0.05$) | +0.2†<br>($\geq 0.05$) | $\leq 0.73$ |
| Pneumonia admission | 69 / 690 | –0.05<br>(–0.22 to +0.11) | –0.17<br>(–0.31 to –0.01) | +0.26<br>(0.005) | +0.27<br>(0.005) | 0.93 |
| Fracture or appendicitis admission | 130 / 1,298 | +0.04<br>(–0.07 to +0.15) | +0.09<br>(–0.02 to +0.19) | –0.02<br>(0.80) | –0.08<br>(0.17) | $>0.99$ |
| Admission, any cause | 859 / 8,247 | +0.02<br>(–0.03 to +0.06) | +0.02<br>(–0.03 to +0.07) | +0.02<br>(0.54) | 0.00<br>(0.86) | $>0.99$ |

**Table S5.**
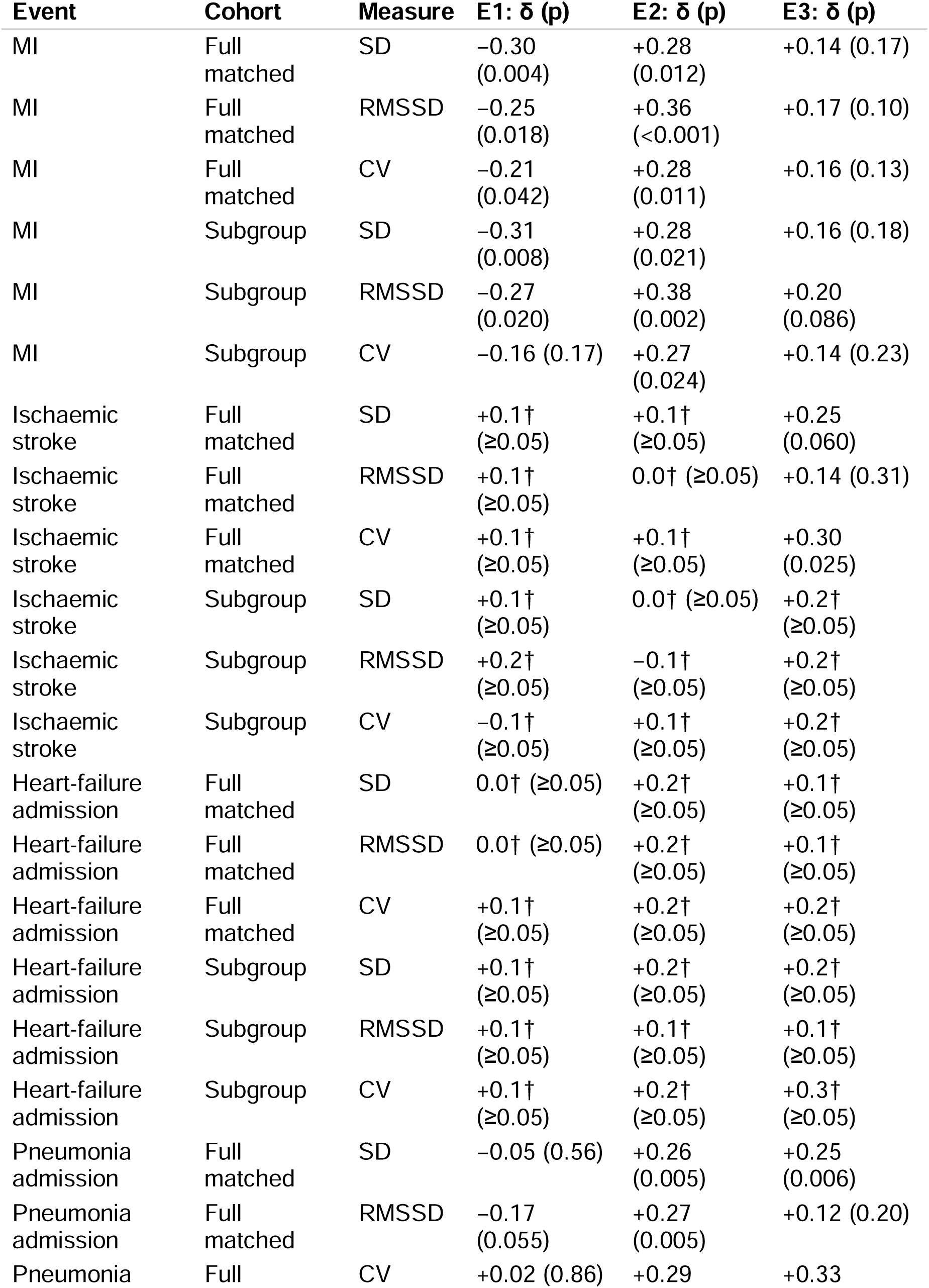
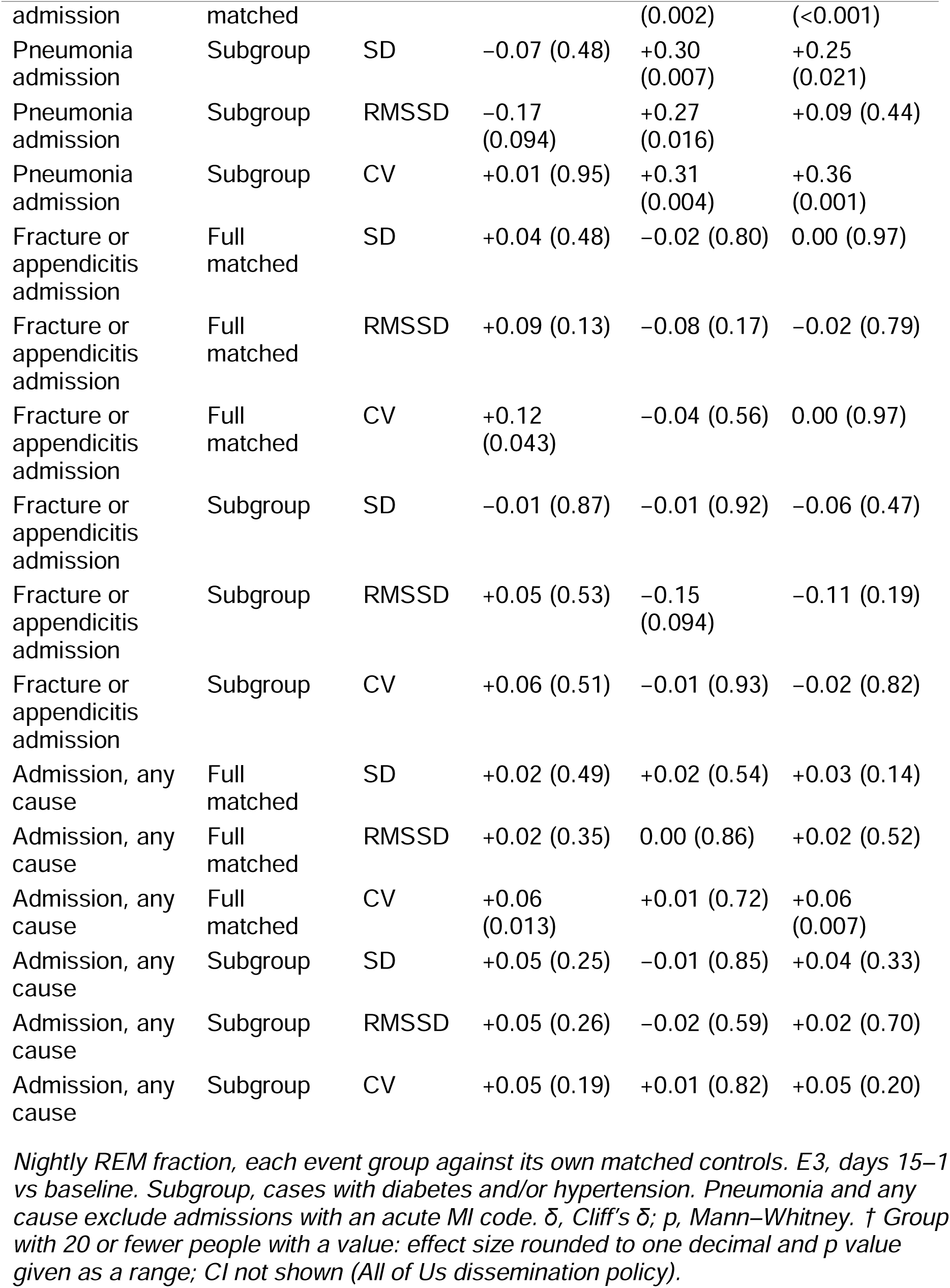
REM-fraction variability before MI and other acute events, all endpoints.

**Table S6.** Matched sets whose case had no recorded symptoms or treatment.

| Measure | Endpoint | Full: $\delta$ (p) | Full: OR per SD (95% CI) | Subgroup: $\delta$ (p) | Subgroup: OR per SD (95% CI) |
| --- | --- | --- | --- | --- | --- |
| REM fraction SD | E1 (d30–16 vs baseline) | –0.30 (0.011) | 0.55 (0.35–0.87) | –0.3† (<0.05) | 0.5† |
| REM fraction SD | E2 (d15–1 vs d30–16) | +0.24 (0.050) | 1.78 (1.09–2.93) | +0.2† ($\geq 0.05$) | 1.8† |
| REM fraction SD | E3 (d15–1 vs baseline) | +0.10 (0.39) | 1.13 (0.74–1.71) | +0.1† ($\geq 0.05$) | 1.2† |
| REM fraction RMSSD | E1 (d30–16 vs baseline) | –0.24 (0.039) | 0.57 (0.36–0.90) | –0.3† (<0.05) | 0.5† |
| REM fraction RMSSD | E2 (d15–1 vs d30–16) | +0.34 (0.006) | 1.99 (1.19–3.31) | +0.4† (<0.05) | 2.2† |
| REM fraction RMSSD | E3 (d15–1 vs baseline) | +0.16 (0.19) | 1.29 (0.86–1.94) | +0.2† ($\geq 0.05$) | 1.4† |
| REM fraction CV | E1 (d30–16 vs baseline) | –0.30 (0.011) | 0.47 (0.29–0.77) | –0.2† ($\geq 0.05$) | 0.5† |
| REM fraction CV | E2 (d15–1 vs d30–16) | +0.24 (0.054) | 1.54 (0.98–2.43) | +0.2† ($\geq 0.05$) | 1.5† |
| REM fraction CV | E3 (d15–1 vs baseline) | +0.06 (0.60) | 1.06 (0.71–1.57) | 0.0† ($\geq 0.05$) | 1.0† |

*Matched sets whose case had no angina (I20.x) or chest-pain (R07.x) code and no newly started beta-blocker, nitrate, antiplatelet or calcium-channel blocker in days 90–1 before the index date. In the subgroup with diabetes and/or hypertension, 20 or fewer cases were asymptomatic. † Group with 20 or fewer people with a value: effect size rounded to one decimal and p value given as a range; CI not shown (All of Us dissemination policy).*

**Table S7.**
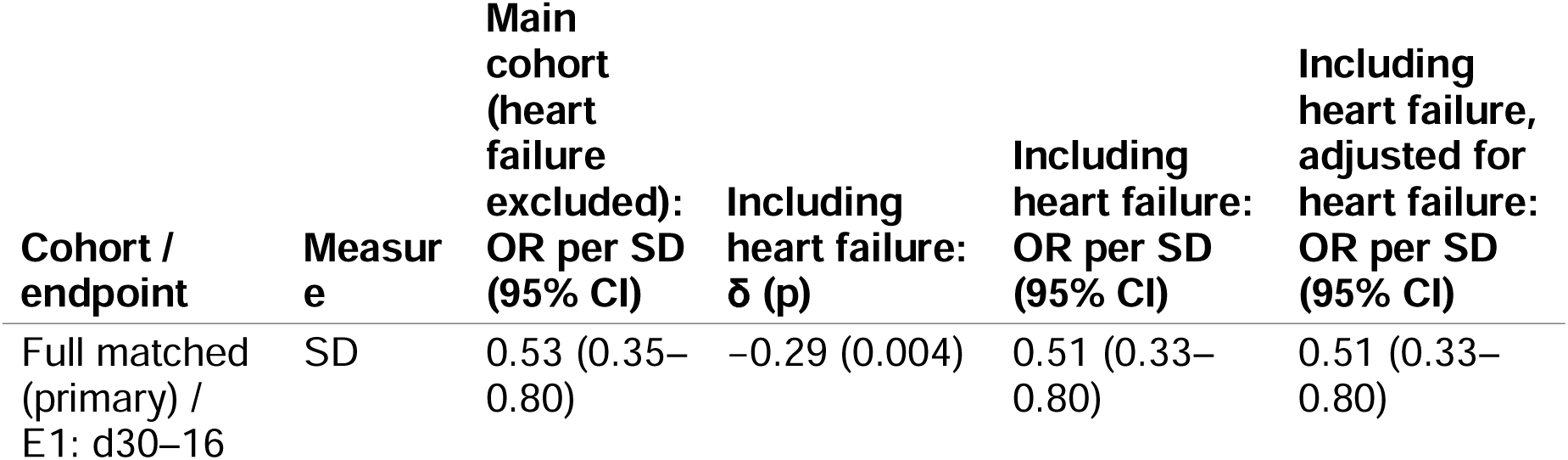
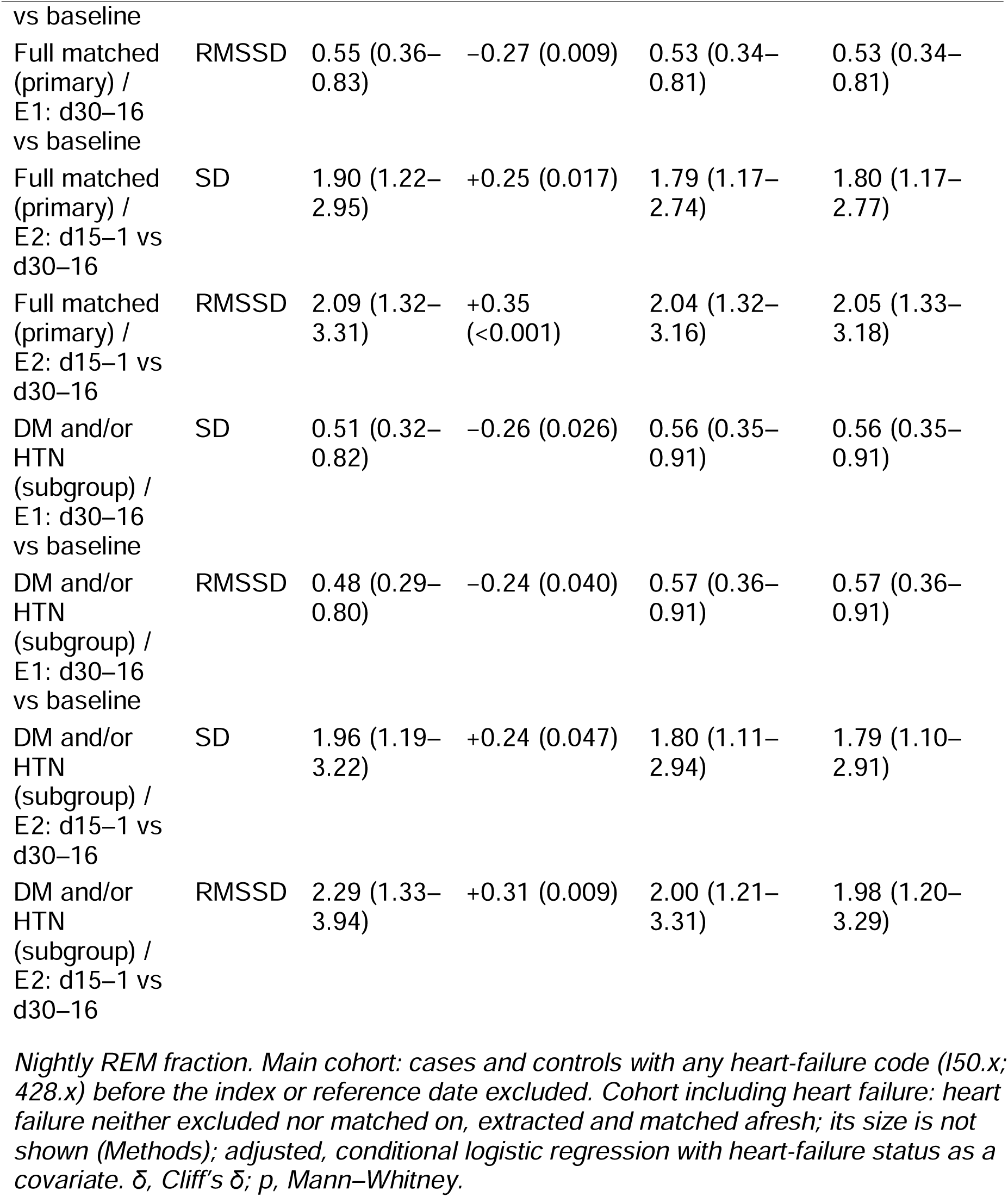
Heart failure: cohort including heart failure, and adjustment for heart failure.

**Table S8.**
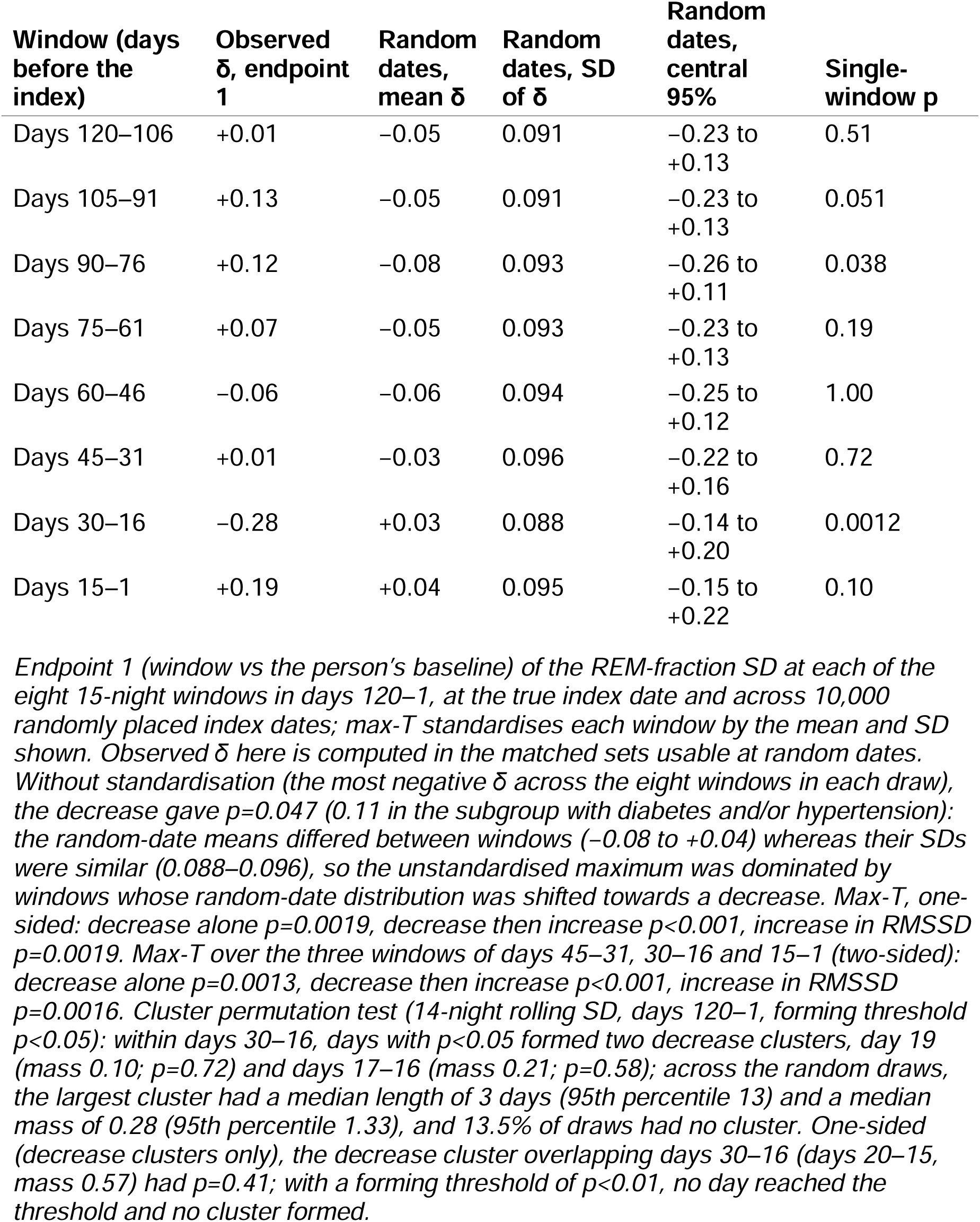
Random index dates: distribution of Cliff’s δ at each window, and the cluster test (full matched cohort, SD)

**Table S9.** Index date at the first angina code.

| Index date | Measure | Full: $\delta$ (p) | Full: OR per SD (95% CI) | Subgroup : $\delta$ (p) | Subgroup: OR per SD (95% CI) |
| --- | --- | --- | --- | --- | --- |
| Original index (MI code) | SD, E1 | -0.30 (0.004) | 0.53 (0.35–0.80) | -0.31 (0.008) | 0.51 (0.32–0.82) |
| Original index (MI code) | RMSSD, E1 | -0.25 (0.018) | 0.55 (0.36–0.83) | -0.27 (0.020) | 0.48 (0.29–0.80) |
| Original index (MI code) | SD, E2 | +0.28 (0.012) | 1.90 (1.22–2.95) | +0.28 (0.021) | 1.96 (1.19–3.22) |
| Original index (MI code) | RMSSD, E2 | +0.36 (<0.001) | 2.09 (1.32–3.31) | +0.38 (0.002) | 2.29 (1.33–3.94) |
| Re-indexed to first angina code (I20.x) | SD, E1 | -0.31 (0.003) | 0.53 (0.35–0.81) | -0.32 (0.007) | 0.52 (0.33–0.83) |
| Re-indexed to first angina code (I20.x) | RMSSD, E1 | -0.28 (0.007) | 0.53 (0.34–0.81) | -0.31 (0.007) | 0.46 (0.27–0.77) |
| Re-indexed to first angina code (I20.x) | SD, E2 | +0.28 (0.011) | 1.91 (1.21–2.99) | +0.28 (0.020) | 1.97 (1.18–3.28) |
| Re-indexed to first angina code (I20.x) | RMSSD, E2 | +0.39 (<0.001) | 2.18 (1.37–3.47) | +0.41 (<0.001) | 2.41 (1.39–4.18) |
| Re-indexed to first angina or chest-pain code (I20.x/R07.x) | SD, E1 | -0.31 (0.003) | 0.53 (0.35–0.81) | -0.32 (0.006) | 0.52 (0.32–0.83) |
| Re-indexed to first angina or chest-pain code (I20.x/R07.x) | RMSSD, E1 | -0.27 (0.010) | 0.54 (0.35–0.83) | -0.30 (0.010) | 0.47 (0.28–0.79) |
| Re-indexed to first angina or chest-pain code (I20.x/R07.x) | SD, E2 | +0.28 (0.012) | 1.88 (1.20–2.94) | +0.28 (0.020) | 1.94 (1.17–3.20) |
| Re-indexed to first angina or chest-pain code (I20.x/R07.x) | RMSSD, E2 | +0.38 (<0.001) | 2.12 (1.34–3.37) | +0.40 (<0.001) | 2.33 (1.35–4.02) |
| Re-indexed sets removed | SD, E1 | -0.32 (0.003) | 0.51 (0.33–0.78) | -0.33 (0.006) | 0.48 (0.29–0.80) |
| Re-indexed sets removed | RMSSD, E1 | -0.29 (0.008) | 0.51 (0.33–0.80) | -0.33 (0.007) | 0.43 (0.25–0.74) |
| Re-indexed sets removed | SD, E2 | +0.24 (0.034) | 1.79 (1.14–2.82) | +0.24 (0.059) | 1.83 (1.10–3.05) |
| Re-indexed sets removed | RMSSD, E2 | +0.35 (0.002) | 2.01 (1.26–3.22) | +0.36 (0.004) | 2.19 (1.25–3.83) |

*Nightly REM fraction. Cases whose first-ever code lay in days 90–1 before the MI index date were re-indexed to that code, and their matched controls' reference dates moved back by the same number of days; all windows were recomputed. The number of re-indexed cases is not shown (20 or fewer).*

