## Supplementary material for "A transient decrease in night-to-night variability of REM sleep before myocardial infarction": Graphical abstract


### Summary

Night-to-night variability of the REM-sleep fraction decreases at days 30–16 before a first myocardial infarction and returns towards baseline over the next two weeks. The decrease was absent at the same time before admissions for pneumonia, fracture or appendicitis and any cause.


### Study design

- Matched case–control study in the All of Us Research Program
- Each person's sleep compared with their own baseline (days 360–121 before the event)

### Population

**44,270**
participants with sleep-stage data and health records

**45/450**
first MI/matched controls

**859**
admissions for any cause, each with matched controls

### Results

REM-sleep variability before the event, relative to matched controls

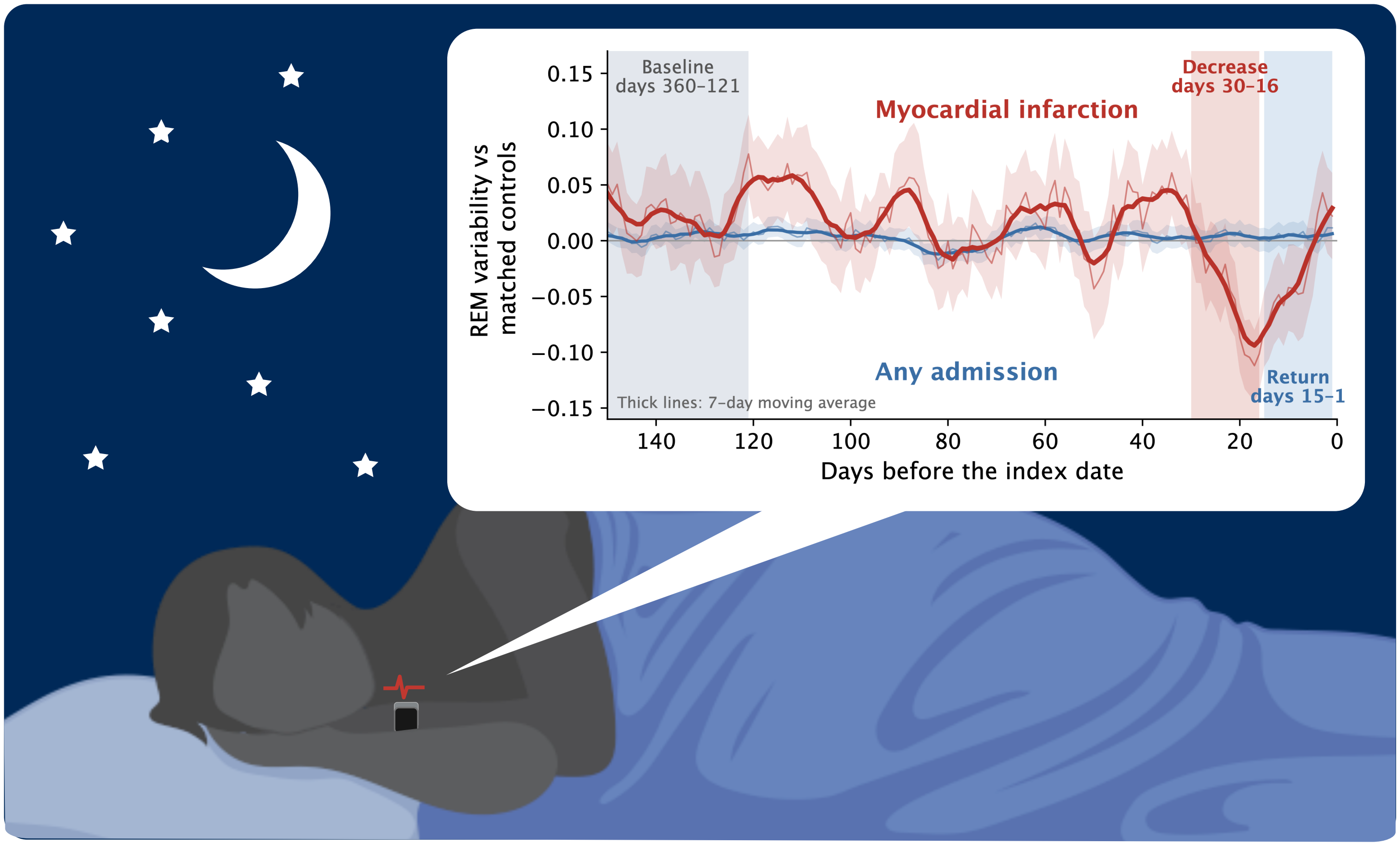


MI • DAYS 30–16 VS BASELINE

**s.d. Cliff's δ −0.30**

*P* = 0.004; OR per s.d. 0.53 (95% CI 0.35–0.80)

ANY ADMISSION • DAYS 30–16

**s.d. Cliff's δ +0.02**

No decrease before admission

CONCLUSION A transient change in REM-sleep variability before MI, detectable against each person's own baseline with a consumer wearable.
